# Polygenic burden of ubiquitin system genes in schizophrenia: focus on prenatal neurodevelopment

**DOI:** 10.64898/2026.07.28.26359147

**Authors:** Gemma Riquelme-Alacid, María Guardiola-Ripoll, Carmen Almodóvar-Payá, Daniel Herrera-Escartín, Noemí Hostalet, Elena Rodriguez-Cano, Raymond Salvador, Salvador Sarró, Amalia Guerrero-Pedraza, Josep Salavert, Llanos Torres, Antonio Arévalo, Mercè Madre, Edith Pomarol-Clotet, Belén Ramos, Mar Fatjó-Vilas

**Author notes:** Senior and corresponding authors and.

## Abstract

Schizophrenia (SZ) is a highly heritable psychiatric disorder with neurodevelopmental origins and a marked impact on cognition. Although alterations in the ubiquitin system have been reported in SZ, the contribution of common genetic variation within this system remains unclear.

Using polygenic scores (PGS) analysis, we assessed the contribution of common SZ-associated variation within ubiquitin system genes (USG) to the disorder susceptibility and whether this contribution varies according to USG spatiotemporal brain expression patterns. We further explored the association of these PGS with cognitive performance.

We defined a Gene Ontology-based panel of 1,450 autosomal USG (global USG panel; gUSG) and tested its enrichment for SZ-associated variation. We calculated the PGS of this panel in 183 individuals with SZ and 127 healthy controls (HC). BrainSpan data were used to stratify the gUSG into different panels by developmental stage (prenatal or postnatal) and brain region (prefrontal cortex and cerebellum). Cognitive evaluation was based on premorbid and current intelligence quotient (IQ), memory and executive function tests.

USG were enriched for SZ-associated variation, and individuals with the disorder showed a higher polygenic burden within this system. The strongest associations involved USG expressed during prenatal development, particularly in the prefrontal cortex. Within SZ, the gUSG-PGS was associated with lower premorbid and current IQ, whereas the prenatal-prefrontal PGS was associated with poorer memory. Together, these findings support a role for USGs in the genetic architecture of SZ and suggest that common variation within this system may link genetic susceptibility to neurodevelopmental processes and cognitive heterogeneity in SZ.

## 1. Introduction

Schizophrenia (SZ) is a complex psychiatric disorder with a lifetime prevalence of approximately 1%, often resulting in significant impairments in daily functioning and reduced quality of life (Kahn et al., 2015; Leucht et al., 2025). Clinically, SZ is characterised by heterogeneous outcomes across individuals and within the same individual over time, including positive and negative symptoms as well as cognitive deficits (Guo et al., 2019; McCutcheon et al., 2023; Sheffield and Barch, 2016).

Multiple studies support the neurodevelopmental hypothesis of SZ (Leucht et al., 2025; Owen et al., 2023; Weinberger, 1987), which posits that the disorder originates from disruptions during critical stages of early brain development. In particular, the prenatal stage is characterised by rapid and dynamic neurodevelopmental processes, including neural proliferation, migration, and early circuit formation, which are tightly regulated at the molecular level and highly sensitive to genetic and environmental perturbations (Kang et al., 2011; Owen et al., 2023). The impact of such perturbations may lead to brain functional and structural abnormalities that disrupt key neurodevelopmental trajectories and manifest later as clinical and cognitive symptoms, particularly in early adulthood (Kahn et al., 2015; Schmidt and Mirnics, 2015).

The role of genetic factors is supported by an estimated heritability of around 80% (Sullivan et al., 2003) and by the identification of several common genetic variants with small effects that collectively increase the risk of developing SZ, as shown in genome-wide association studies (GWAS; Ripke et al., 2014; Trubetskoy et al., 2022). These findings have enabled the use of quantitative measures such as polygenic scores (PGS), which, based on GWAS data, estimate an individual’s cumulative genetic burden for that given trait or a disorder (Wray et al., 2014).

Functional analyses of GWAS data have shown that SZ-associated variants tend to cluster in genes involved in neurodevelopmental processes, including neuronal differentiation, maturation and synaptic plasticity (Ripke et al., 2014; Trubetskoy et al., 2022). These genetic influences may contribute to the broad molecular heterogeneity observed in SZ, including abnormalities in gene expression and protein homeostasis (Andrews et al., 2017; Bousman et al., 2019; Nucifora et al., 2019), thereby affecting biological systems relevant to SZ pathophysiology (Arion et al., 2015). Among these processes, proteostasis has emerged as a key mechanism for maintaining cellular function, as it coordinates protein synthesis, folding and degradation (Hipp et al., 2019). This regulation is particularly critical during neurodevelopment, synaptic plasticity and circuit stabilization, where precise temporal and spatial control of protein abundance is required (Grochowska et al., 2022).

The ubiquitin system is one of the main regulators of proteostasis and participates in the selective degradation of proteins and the maintenance of cellular function (Franić et al., 2021; Pohl and Dikic, 2019). The ubiquitin system comprises E1 ubiquitin-activating enzymes, E2 ubiquitin-conjugating enzymes, E3 ubiquitin ligases and deubiquitinating enzymes, which dynamically and reversibly regulate the ubiquitination of target proteins, thereby modulating their localisation, function or degradation (Clague et al., 2019; Damgaard, 2021; Oh et al., 2018; Yang et al., 2021). Overall, the ubiquitin system constitutes a highly complex and versatile regulatory network, crucial for a wide range of processes related to brain development and maturation (Ambrozkiewicz and Kawabe, 2015; Folci et al., 2020; Kawabe and Brose, 2011; Zajicek and Yao, 2021). In this regard, several components of the ubiquitin system show high expression during prenatal stages, consistent with a key role in early neurodevelopmental events such as neuronal differentiation, migration, and synaptic assembly (Hale and Bashaw, 2025; Magnati et al., 2024). Its relevance has also been demonstrated in several psychiatric disorders, where disruptions in ubiquitin-dependent processes have been associated with abnormal protein accumulation, impaired synaptic remodelling, altered neurodevelopmental trajectories, and dysregulated cellular stress responses (Andrews et al., 2017; Bousman et al., 2019; Hegde et al., 2025; Meiklejohn et al., 2019; Nakatochi et al., 2025; Rubio et al., 2013). In the context of SZ, altered expression and activity of components of this system have been reported in brain regions such as the prefrontal cortex (PFC) as well as in peripheral blood, suggesting their potential role as biomarkers (Andrews et al., 2017; Bousman et al., 2019; Meiklejohn et al., 2019).

Despite the growing interest in their functional role, the contribution of genetic variability in ubiquitin system genes (USG) to SZ risk has been underexplored. Some studies have identified Single Nucleotide Polymorphisms (SNPs) in USG associated with increased vulnerability for this disorder (Liu et al., 2017; Middleton et al., 2002; Trubetskoy et al., 2022); however, no study has investigated whether the polygenic burden in USG could provide relevant insights into SZ susceptibility mechanisms.

According to all the above-mentioned, the present study aimed, first, to explore whether common genetic variation in USG, quantified using PGS, is associated with susceptibility for SZ. Second, we examined whether this association is influenced by the spatiotemporal expression patterns of USG during brain development. Finally, we assessed whether genetic burden in USG is related to the cognitive deficits observed in SZ. Together, this multi-level approach is designed to uncover novel insights into the functional relevance of the ubiquitin system in SZ pathophysiology.

## 2. Materials and Methods

### 2.1 Participants

A total of 352 participants were initially recruited for this study. Patients were recruited from inpatient and outpatient units of Fundació Hospitalàries Sant Boi (Barcelona, Spain) and healthy controls (HC) were recruited from the same geographical area. All participants were of European descent, between 18 and 65 years old. All met the same exclusion criteria, which included suffering from major medical illness, conditions affecting cognitive or brain function, neurological conditions, history of head trauma with loss of consciousness and current or past substance abuse or dependence. Additionally, for HC, exclusion criteria also included personal or family history of psychiatric service contact or treatment.

SZ diagnosis was confirmed using DSM-IV-TR criteria through structured interviews with two psychiatrists. Clinical symptoms of SZ patients were assessed using the Positive and Negative Syndrome Scale (PANSS) (Kay et al., 1987), which derives three scores: positive, negative and general psychopathology symptoms scores.

After genetic quality control, 42 individuals were excluded for not meeting the predefined criteria (see section 2.3). The final sample for genetic analyses comprised 310 participants: 183 individuals diagnosed with SZ (51 females and 132 males; mean age (SD) = 42.9 (11.7) years) and 127 HC (62 females and 65 males; mean age (SD) = 38.1 (11.8) years). For the cognitive analyses, participants were selected based on the availability of cognitive data. Those with an estimated premorbid intelligence quotient (IQ) below 70 were excluded, and groups were matched for age and sex. The resulting subsample compromised 242 participants: 130 individuals with SZ (43 females and 87 males; mean age (SD) = 41.3 (10.7) years) and 112 HC (51 female and 61 males; mean age (SD) = 38.9 (11.8) years).

### 2.2 Cognitive Assessment

Cognitive performance was characterised using four standardised neuropsychological instruments. Premorbid IQ was estimated using the Spanish version of the Word Accentuation Test (Test de Acentuación de Palabras; TAP) (Gomar et al., 2011), a reading test in which participants are required to pronounce written words without accent marks. Raw TAP scores range from 0 to 30 and were converted into estimated IQ scores (Gomar et al., 2011). Participants with TAP-derived IQ scores below 70 were excluded from subsequent analyses. Current IQ was estimated using four subtests of the Wechsler Adult Intelligence Scale III (WAIS-III) (Wechsler, 1997): vocabulary, similarities, block design and matrix reasoning. Executive functioning was assessed using the Behavioural Assessment of the Dysexecutive Syndrome (BADS) (Wilson et al., 1996), which provides a global score derived from six tasks designed to reflect executive functioning in everyday life. Memory performance was measured using the Wechsler Memory Scale (WMS) (Wechsler, 1945), through a composite score that includes results from four subtests: logical memory, face recognition, digit span, and letter-number sequencing.

### 2.3 Genomic DNA extraction, genotyping, quality control, and imputation

Genomic DNA was extracted and genotyped following standard procedures. After quality control, phasing, and genotype imputation, 7.6 million variants were retained. Detailed protocols and filtering criteria are provided in Supplementary Methods 1.

### 2.4 Selection of ubiquitin system genes

USG were selected using a Gene Ontology (GO)-based strategy implemented in R (Supplementary Figure S1). All GO terms containing the word “ubiquitin” were retrieved. To increase specificity, terms including “ubiquitin-like”, “sumo”, “NEDD”, “UFM”, “no ubiquitin”, and “ubiquitin independent” were excluded, resulting in a final set of 250 GO terms (Supplementary Table 1).

Finally, a total of 1,450 autosomal human genes were retained, forming the global ubiquitin system genes panel (gUSG) (Supplementary Table 2a). For further details on this procedure, see Supplementary Methods 2.

### 2.5 Competitive gene-set analysis of ubiquitin system genes

To evaluate whether the USG showed stronger associations with SZ than others genes, we performed a competitive gene-set analysis (GSA) using MAGMA v1.10 (de Leeuw et al., 2015) and the summary statistics from the SZ GWAS by Trubetskoy et al., (2022). Gene level association statistics were derived from GWAS SNPs mapped to genes and used to test the competitive enrichment of the gUSG panel (n = 1,450) relative to the rest of the genome. To determine whether the observed enrichment exceeded chance expectations, 1,000 matched random gene sets were generated and analysed using the same pipeline, yielding an empirical p-value based on the resulting null distribution. For further details on this procedure, see Supplementary Methods 3.

### 2.6 Spatiotemporal characterisation and tissue association of the ubiquitin system genes panel

To guide downstream analyses, we explored the temporal expression of gUSG, and the tissue-specific enrichment of SZ-associated variants located within these genes. These exploratory in silico analyses were performed using the FUMA platforms, and they are described in detail in Supplementary Methods 4. Key observations relevant for downstream analyses showed, first, that genes in the gUSG panel (n = 1,450) tend to exhibit lower relative expression compared with the genome-wide average during early childhood and middle adulthood (supplementary Figure S2), indicating the differential expression of ubiquitin-related genes after birth (Magnati et al., 2024). Second, tissue enrichment analysis of SZ-associated SNPs located in gUSG genes revealed a tissue-specific expression profile enriched in cerebellar (p = 4.6 x 10⁻⁵) and prefrontal areas (frontal cortex BA9: p = 1.0 x 10^-3^; brain cortex: p = 1.9 x 10^-3^) consistent with prior studies (Andrews et al., 2017; Hwang et al., 2022) (supplementary Figure S3).

### 2.7 Selection of USG panels based on spatiotemporal expression in brain development

To examine whether the impact of USG on SZ vulnerability may be modulated by their spatiotemporal expression, we generated six gene subsets from the gUSG list based on transcriptomic data from the Atlas of the Developing Human Brain (BrainSpan, www.brainspan.org) (Kang et al., 2011; Sunkin et al., 2013). We first defined two panels of genes overexpressed in either prenatal or postnatal stage across the brain: the prenatal USG (preUSG) and the postnatal USG (postUSG). Then, within these panels, we defined 4 panels based on the brain regions identified in the enrichment analysis (prefrontal cortex, PFC; and cerebellum, CB): 2 prenatal panels prePFC_USG and preCB_USG) and 2 postnatal panels (postPFC_USG and postCB_USG). These six panels, along with the gUSG, were subsequently used to calculate PGS and explore their associations with SZ and cognitive traits.

Full details of this section procedure are provided in Supplementary Methods 5 and Supplementary Tables 3a-3f.

### 2.8 Polygenic score estimation

Using the summary statistics from the European subsample of the 2022 GWAS in SZ (Trubetskoy et al., 2022), the seven panel-specific PGS were calculated using PLINK 1.9 (Chang et al., 2015) with the clumping and thresholding (C+T) approach (Privé et al., 2019). First, linkage disequilibrium (LD) filtering was applied using the European population from the 1000 Genomes Project Phase 3 reference panel as a baseline. An r² > 0.5 threshold and 250 kb windows were used, prioritising variants with the highest statistical significance (see Supplementary Tables 4a-4g). We applied 13 p-values thresholds, from 5x10^-8^ to 1.0. PGS were calculated as the sum of risk alleles weighted by GWAS effect size. Additionally, a global PGS (PGS_globalSZ) was calculated using the same procedure as the panels based on PGSs-GSUs but including all SNPs from the reference GWAS (Trubetskoy et al., 2022).

### 2.9 Statistics

Statistical analyses were performed using SPSS (version 24.0; IBM Corp., Armonk, NY, USA) and RStudio (version 4.5.0; (R Studio Team, 2021; RStudio, 2011)). Visualisations were generated with the ggplot2 library (Wickham et al., 2016).

#### 2.9.1 Polygenic score selection and association with schizophrenia diagnosis

First, each threshold was independently tested using unadjusted binary logistic regression solely for optimal threshold selection. The threshold yielding the lowest p-value and highest Nagelkerke pseudo-R^2^ was then carried forward to subsequent analyses (supplementary figure S4). Then, to analyse the associations between USG polygenic burden and SZ status, we fitted a separate adjusted binary logistic regression (BLR) model for each panel using its selected optimal threshold. Each model was constructed hierarchically in two blocks: Block1 included age, sex, and the first four principal ancestry components, while Block2 incorporated the corresponding PGS. The contribution of the PGS to the model was assessed both by its statistical significance as a predictor and by the increase in Nagelkerke’s pseudo-R² (explained variance) in Block2.

The discriminative capacity of the models was evaluated using receiver operating characteristic (ROC) curves, employing the caret (Max, 2008) and pROC (Robin et al., 2011) packages in RStudio (v4.5.0). The PGS_globalSZ was included as reference for comparative purposes in predictive model validation analyses. Stratified cross-validation was applied using a k-fold scheme (5 folds and 5 repetitions), using a fixed randomisation seed (using the set.seed(123) function in r). Sensitivity, specificity, area under the curve (AUC), and the corresponding 95% confidence interval (CI) were estimated for each PGS associated with the different USG panels. Finally, differences in the discriminative capacity of the models were compared using DeLong’s test for correlated ROC curves, assuming that non-significant differences between the ROC curves indicate comparable predictive utility.

#### 2.9.2 Association between ubiquitin system genes-derived polygenic risk score and cognitive functioning

Multiple linear regression (MLR) models were used to assess whether PGS modulated cognitive performance. The dependent variables were the scores obtained from the neuropsychological scales employed: TAP, IQ, BADS, WMS. Each model was structured into two blocks: Block1 included control variables (age, sex, diagnosis, and the first four ancestry components), and Block2 incorporated the corresponding PGS.

Additionally, we performed stratified analyses by diagnostic group (HC and SZ) to explore potential differential patterns. In HC models, Block1 included age, sex, and the first four ancestry components, while in SZ models, chlorpromazine equivalent dose (CPZ) and illness duration (years since diagnosis) were also incorporated. In all cases, Block2 included the corresponding PGS.

#### 2.9.3 Multiple comparison adjustment

All results were adjusted for multiple comparisons using the Benjamini-Hochberg procedure, with statistical significance defined as an FDR-adjusted p-value (pFDR) < 0.05.

## 3. Results

### 3.1 Sociodemographic, Clinical, and Cognitive Characteristics of Participants

The sociodemographic, clinical, and cognitive characteristics of the cognition subsample are summarised by diagnostic group in Table 1. Since, it was matched for age and sex, no significant differences were observed between groups. Clinical symptoms scores are reported for descriptive purposes. Across all cognitive measures, HC scored significantly higher than SZ group.

**Table 1.**
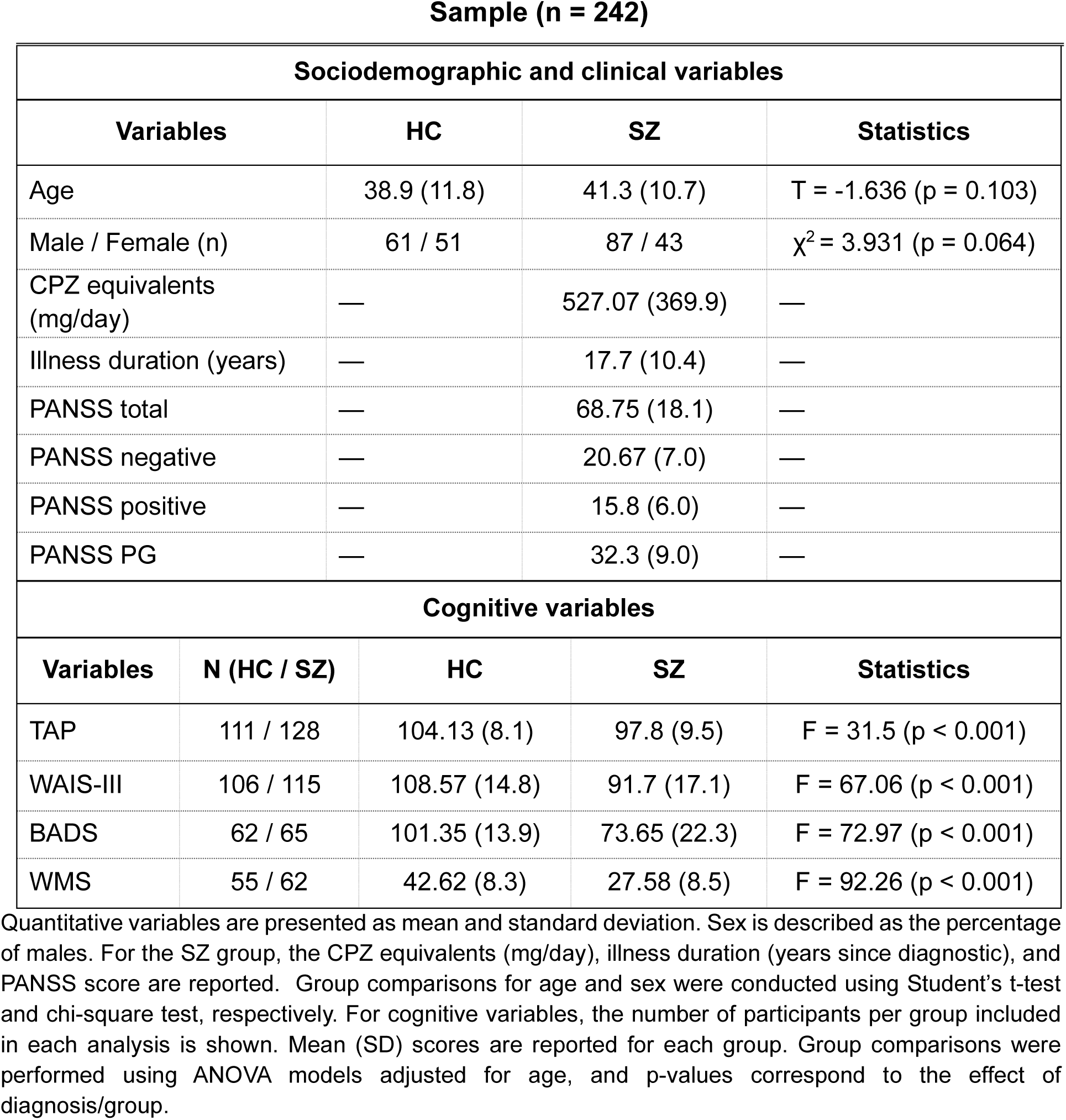
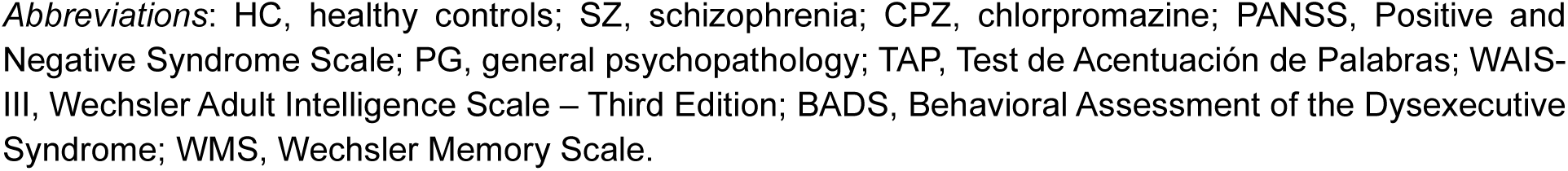
Sociodemographic, clinic and cognitive variables.

### 3.2 Gene set analysis of ubiquitin system genes

Competitive GSA showed that the gUSG panel was significantly enriched for SZ-associated variants (β = 0.080, SE = 0.032, p = 0.006). The observed effect exceeded chance expectation relative to 1,000 matched random gene sets (empirical p = 0.002; Figure Supplementary S5).

### 3.3 Association of polygenic burden of ubiquitin system genes with schizophrenia

The RLB models were implemented sequentially. First, none of the adjustment variables included in Block1 were associated with the diagnosis, and together they explained a small proportion of variance (Nagelkerke’s pseudo-R² = 0.070). Second, the inclusion of each PGS in Block2 significantly improved model fit in all cases, except when including those PGS based on cerebellar expression (Table 2). Specifically, the inclusion of PGSs based on gUSG, preUSG, and prePFC_USG panels, increased more than two-fold the explained variance of the models. Next, to evaluate the discriminative capacity of these models, ROC curve analyses were conducted. Previously we had estimated the effect of PGS_globalSZ in our sample (OR=2.06, 95% CI=1.59-2.66; p=3.39x10^-8^) and the ROC curve for this model. The gUSG model performed similarly to the PGS_globalSZ (p=0.446), suggesting that gUSG captures a substantial proportion of SZ-related polygenic signal (Supplementary Figure S6A). When considering the prenatal panels, comparisons revealed no significant difference in AUC between gUSG and the preUSG or prePFC_USG panels, while the preCB_USG model showed lower discriminative accuracy (Supplementary Figure S6B). In contrast, the postnatal panel showed consistently lower AUC values, with limited evidence of added discriminative values (Supplementary Figure S6C).

Taken together, these findings indicate that PGS derived from the gUSG, preUSG, and prePFC_USG panels exhibit the best diagnostic association and model fit. To streamline subsequent analyses and reduce the risk of overfitting, we focused on these three panels in the cognitive analyses presented below.

**Table 2.**
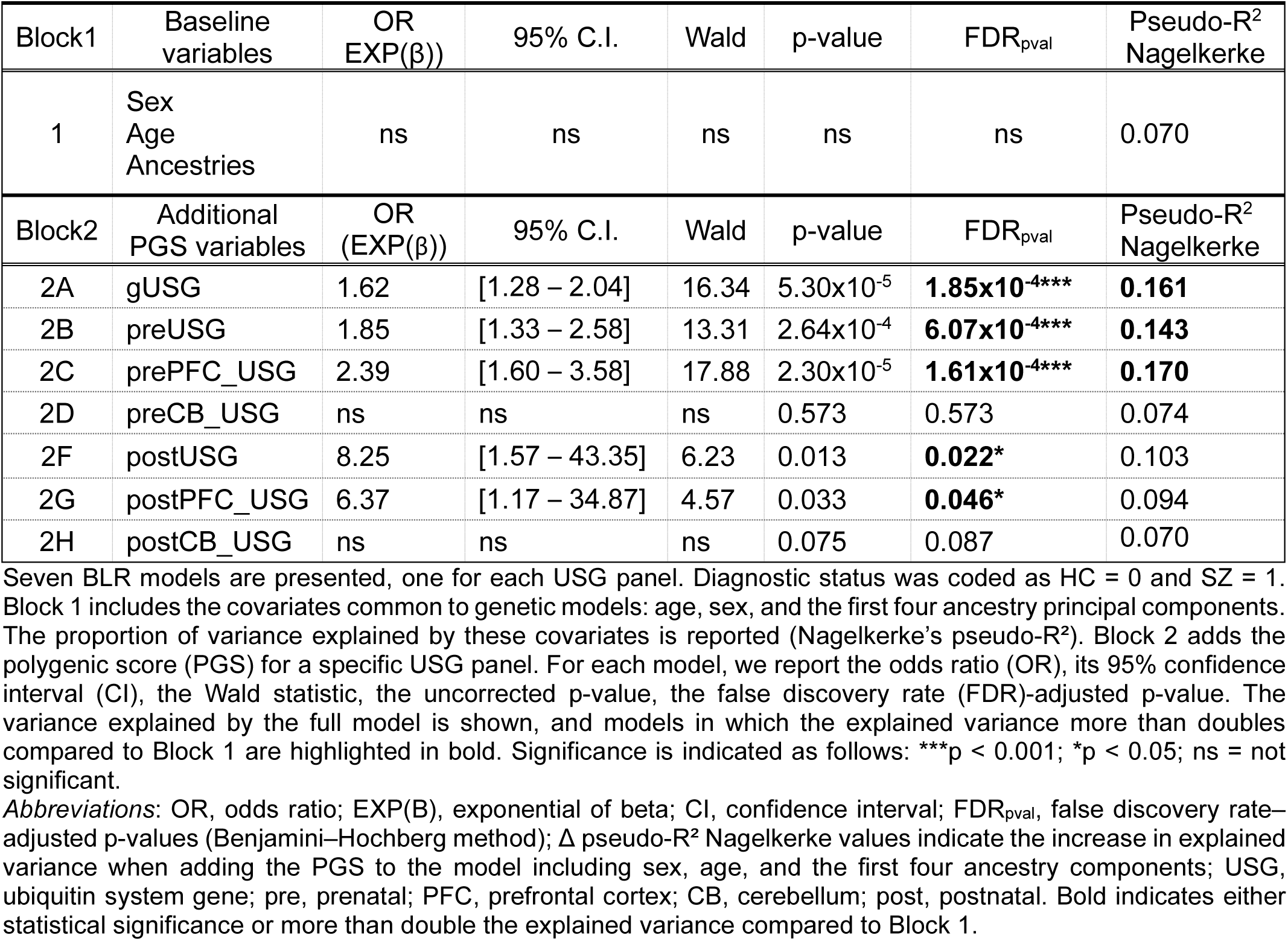
Binary logistic regression (BLR) for each model USG panel.

### 3.4 Association between polygenic load and cognition

We next explored the association between USG-derived PGS and cognitive performance. In the cognitive sample, PGS from the gUSG, preUSG, and prePFC_USG panels were significantly associated with current IQ (WAIS-III) and executive function (BADS), after FDR correction (Table 3). No significant associations were found with premorbid IQ (TAP) or memory performance (WMS) (Supplementary Table 5).

In stratified analyses by diagnostic group, significant associations were found only in the SZ group. In this group, gUSG PGS was associated with both premorbid and current IQ, while prePFC_USG PGS showed associations with memory performance (Table 4). No significant associations were observed in models involving executive functions. All non-significant results are reported in Supplementary Table S6.

**Table 3.**
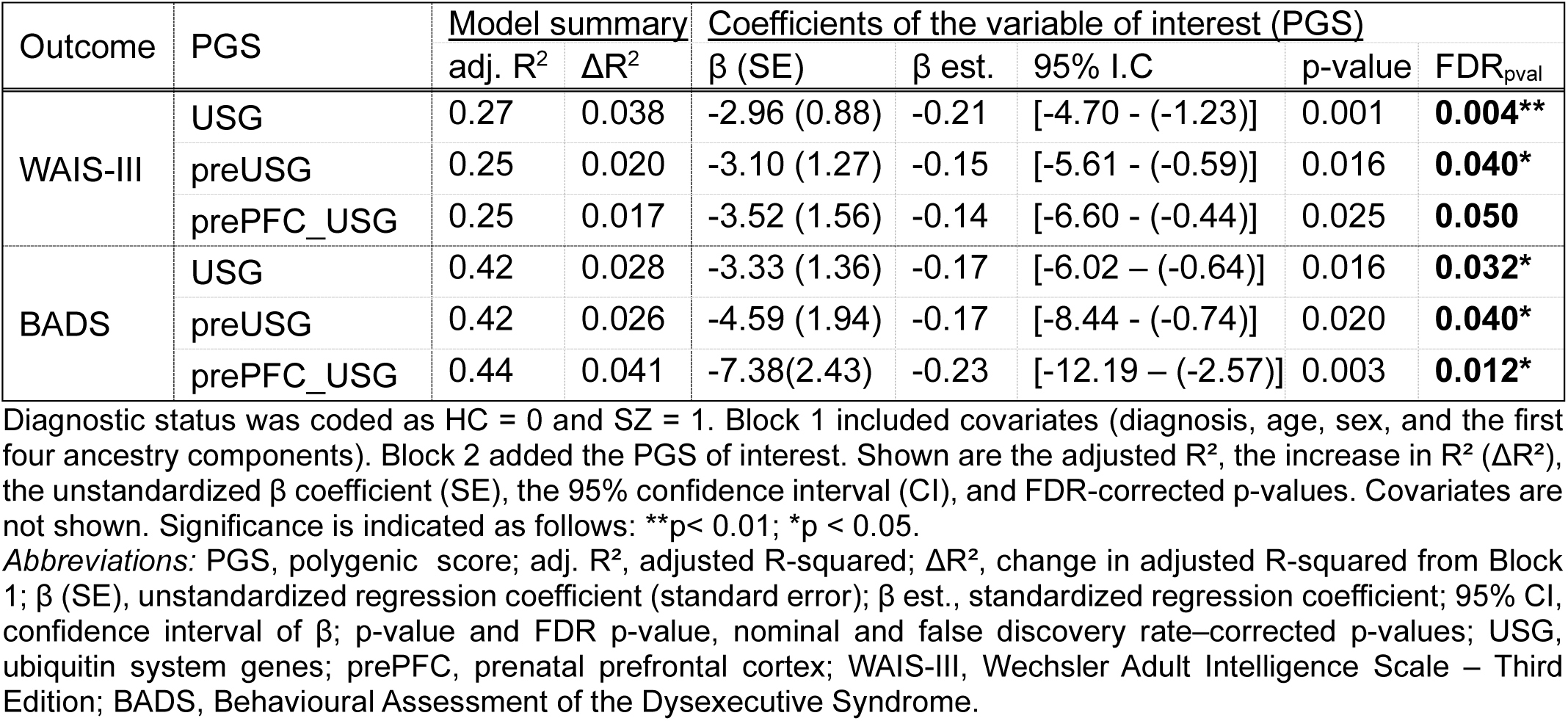
Cognitive outcomes significantly predicted by USG-derived PGS.

**Table 4.**
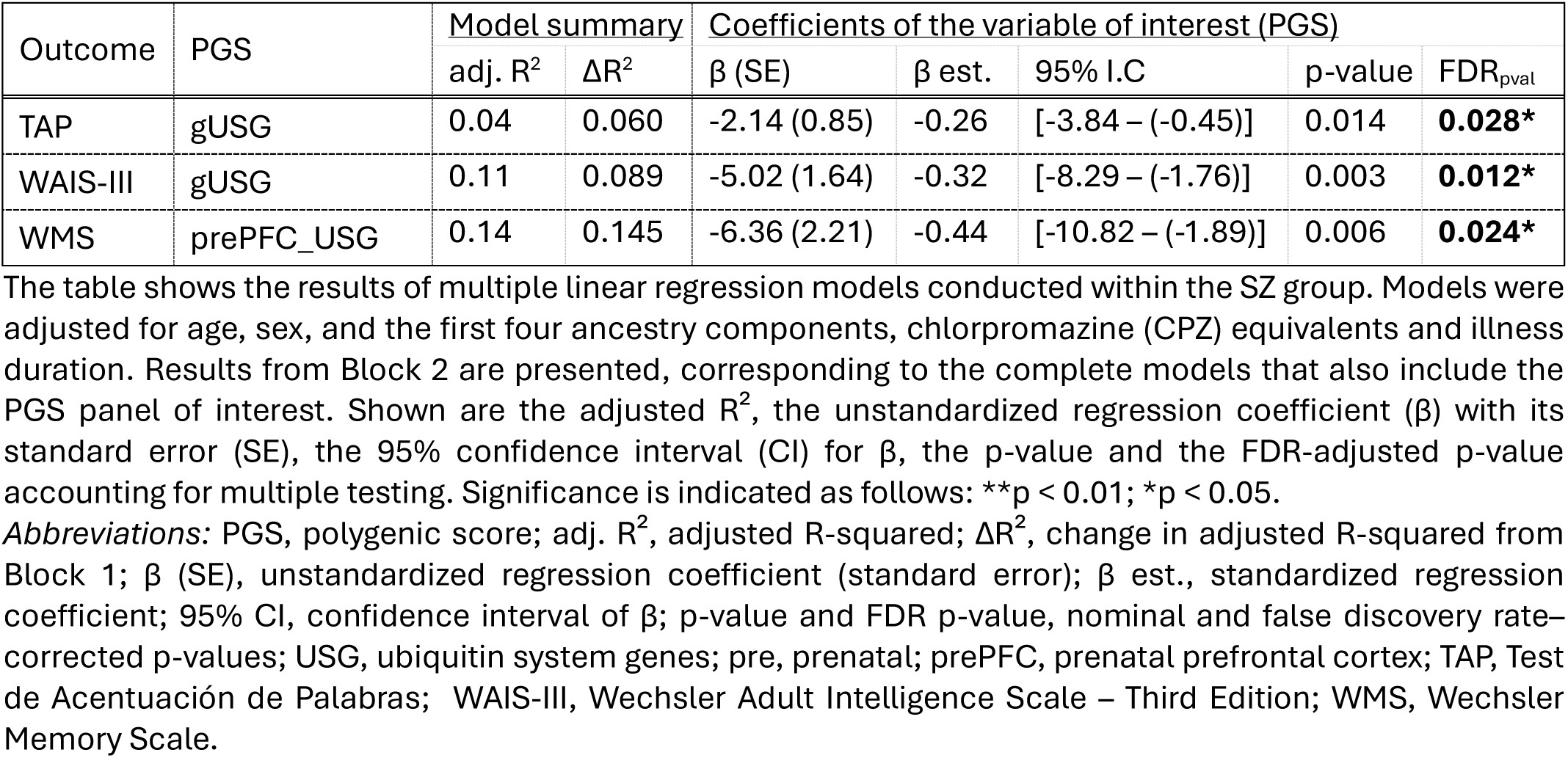
Significant PGS associations with cognitive performance in the schizophrenia group.

## 4. Discussion

This study provides new evidence regarding the contribution of the ubiquitination system to susceptibility to SZ. Through the analysis of common genetic variation in ubiquitination-related genes, we highlight three main findings: 1) individuals with SZ show a higher polygenic load in USGs; 2) this effect is particularly evident for USGs that are overexpressed during prenatal development and in cortical regions; and 3) the higher genetic load in USGs is associated with poorer cognitive performance.

First, we evaluated the contribution of USGs to the polygenic architecture of SZ (Owen et al., 2023; Trubetskoy et al., 2022), offering a new perspective on the involvement of the ubiquitination system in this disorder (Chen et al., 2024; Hegde et al., 2025; Luza et al., 2020). To this end, we applied an approach based on a USG panel, including gene set analysis (GSA), individual PGS, and discriminative capacity (ROC). Our results indicate a significant enrichment of the gUSG panel in the competitive GSA, suggesting that this set of genes captures a meaningful proportion of the genetic signal previously identified through GWAS for SZ risk (Trubetskoy et al., 2022). Furthermore, we observed that the SZ group had a higher polygenic load in USGs than the control group, reinforcing their involvement in the disorder’s susceptibility and suggesting that PGS based on USGs represents a genetic risk marker. Likewise, the gUSG panel showed a discriminative capacity not statistically different to that of the global SZ PGS (Trubetskoy et al., 2022), further supporting the relevance of USGs within the polygenic architecture of the disease. Overall, our results are consistent with previous evidence implicating the ubiquitination system in this disorder, both in studies focused on specific genes (Chen et al., 2008; Cheng et al., 2024) and in studies based on GWAS data. Despite using complementary network and pathway based approaches, these studies converged in identifying ubiquitin-related processes as associated with SZ across independent dataset and ancestries (Chang et al., 2015; Liu et al., 2017). Building on this evidence, our study is, to our knowledge, the first specifically designed to evaluate common genetic variation across a comprehensive USG panel, assessing both gene-set enrichment and individual level polygenic burden in SZ.

This system-level genetic evidence is especially relevant when considered in the context of molecular alterations previously described in SZ. At the transcriptomic level, differential expression patterns in the ubiquitin–proteasome system have been reported between patients with SZ and controls, both in cortical brain regions and peripheral blood samples, leading to the proposal of these alterations as a potential differential molecular signature (Arion et al., 2015; Middleton et al., 2002; Dor and Hertzberg, 2024)). Remarkably, some of these studies have also identified specific ubiquitin expression profiles that can define subgroups related to the severity of the disorder (Dor and Hertzberg, 2024) and schizoaffective symptomatology (Arion et al., 2015).

Proteomic studies further support the role of this system by reporting the accumulation of ubiquitinated proteins in cortical regions, olfactory neurons and blood in patients compared with controls (Bousman et al., 2019; Nucifora et al., 2025, 2019; Rubio et al., 2013). The authors interpret these findings as reflecting both failures in protein degradation and changes in ubiquitination patterns that redirect proteins toward non-degradative pathways. Along the same lines, other studies have suggested that disruption of the ubiquitination system in SZ could have broader molecular effects by altering the regulation of proteins involved in cellular processes and signalling pathways relevant to the disorder (Kliza and Husnjak, 2020; Yalla et al., 2018). Based on this, we hypothesise that the accumulation of genetic risk variability in USGs could be a marker of a compromised balance between ubiquitination and proteostasis, with repercussions for other biological pathways, thus constituting a possible link between the genetic signal and part of the molecular dysfunction observed in SZ. However, further integrative approaches combining genetic, transcriptomic, and proteomic data are needed to enable a more comprehensive characterisation of how variation in USGs translates into functional alterations across molecular layers in SZ.

Next, given the role of the ubiquitination system in key neurodevelopmental processes, ranging from neuronal differentiation and organisation to connectivity and synaptic remodelling (Xue et al., 2025; Zajicek and Yao, 2021), we investigated whether the genetic contribution of USGs is uniformly distributed across developmental stages and brain regions. Our results indicate that this effect is not homogeneous but rather is concentrated in the prenatal period and in the prefrontal cortex. This pattern suggests that genetic variability in USGs could early influence a suboptimal organisation of cortical circuits, particularly prefrontal ones, with persistent effects during their maturation and functioning. This interpretation is consistent with previous studies showing that genes involved in SZ risk tend to exhibit differential expression both in the placenta and during prenatal neurodevelopment (Birnbaum and Weinberger, 2024; Ursini et al., 2021). These authors propose that such expression patterns may confer a biological vulnerability that interacts dynamically with later brain maturation processes, when the corresponding neural circuits complete their development during adolescence and adulthood. In line with this, in a murine neurodevelopmental model of SZ, Andrews et al. (2017) observed dynamic alterations in ubiquitin-proteosome system proteins in the prefrontal cortex, spanning from prenatal stages through adulthood, supporting the notion that this brain region may be particularly vulnerable to early alterations in protein regulation.

On the other hand, our results also showed an association between postnatal USGs and SZ risk, although their lower discriminative capacity suggests a more modest or less consistent contribution to the disorder. Similarly, the signal observed in the cerebellum in the gene enrichment analyses was not supported by individual-level analyses. This discrepancy could reflect the smaller number of genes included in these postnatal and cerebellum panels, which limits statistical power. Also, it could indicate that enrichment and polygenic approaches are capturing different dimensions of the genetic signal (de Leeuw et al., 2015; Wray et al., 2018). Taken together, these findings underscore the need for integrative multilevel approaches to better understand how the spatiotemporal organisation of gene expression modulates schizophrenia vulnerability.

Finally, we evaluated the impact of USGs polygenic burden on cognitive performance, based on the role of the ubiquitination system in key processes for neurocognitive functioning, such as homeostasis and synaptic plasticity (Mabb and Ehlers, 2010; Yang et al., 2024). Our results show that a higher USG polygenic load is associated with poorer cognitive performance, suggesting that genetic variation in this system may influence the biological mechanisms underlying cognitive impairments in SZ. This finding aligns with recent studies linking overall SZ polygenic burden to cognitive deficits (Rosenqvist et al., 2026) and our results extend this evidence by indicating that USGs may account for part of this association. In our cohort, this effect was primarily observed in general cognitive ability and executive functions. Although these domains depend on complex neural networks, their functioning relies on processes of synaptic plasticity and dendritic stability regulated by the ubiquitination system (Hamilton and Zito, 2013; Patrick et al., 2023).

Stratified analyses further showed that the relationship between USGs and cognitive performance was limited to the SZ group, suggesting that the effect of this genetic load on cognition may manifest primarily in the context of the disorder. When we examined the results by panel, we observed that the global panel was associated with current and premorbid IQ, whereas prenatal prefrontal cortex panel showed a more specific relationship with memory. While caution is required due to the limited sample size in stratified analyses, these findings suggest that depending on their temporospatial expression, different subsets of USGs may contribute to cognitive performance. On the one hand, the association of the global panel with both measures of intellectual ability may reflect a broader involvement of the ubiquitination system in regulating proteostasis, which is necessary for maintaining neuronal and synaptic architecture. By contrast, the specific association between the prenatal PFC panel and memory may reflect a genetic vulnerability involving genes active during early PFC development, whose cognitive consequences emerge later through circuit maturation and synaptic plasticity, potentially shaped by subsequent developmental and environmental influences (Ambrozkiewicz and Lorenz, 2025; Hegde, 2017). This interpretation is consistent with evidence linking dysfunction of the ubiquitination system both to alterations in neuronal development and organisation described in mental disorders also characterised by cognitive impairments, such as autism spectrum disorder and intellectual disability (Folci et al., 2020; Kasherman et al., 2020), and to impairments in proteostasis regulation and synaptic plasticity associated with memory alteration in Alzheimer’s disease (Hegde et al., 2019). Taken together, these findings open the door to exploring whether USGs’ variability may contribute to cross-disorder mechanisms underlying shared cognitive traits.

Although we provide new data supporting an association between the ubiquitination system and SZ, our results should be interpreted in light of several limitations that point to future research directions. First, the sample size, particularly in cognitive analyses, may have limited the statistical power to detect small effects. Moreover, the low proportion of female participants precluded a robust assessment of sex-by-PGS interactions. Likewise, the unequal number of genes across panels may have introduced heterogeneity in statistical power, with smaller panels being less sensitive to detect polygenic signal. In addition, the samples used for PGS threshold selection and hypothesis testing overlapped, which may have inflated estimates of model performance. Because our primary aim was to test whether SZ was associated with increased USG polygenic burden rather than to estimates of PGS performance. Second, our analysis focused exclusively on the functional core of the ubiquitination system, excluding ubiquitin-like proteins. Future studies with more inclusive approaches to the ubiquitination system and its interactors could offer a complementary view of how global dysregulation of proteostasis contributes to SZ. Third, another factor that should be considered is the potential effect of antipsychotic treatment on cognitive status. While we accounted for antipsychotic use in the within-patient analyses, its influence cannot be fully excluded. However, it is notable that the evidence indicates that the ubiquitination system is not directly modulated by antipsychotic treatment (Andrews et al., 2017; Luza et al., 2020; Nucifora et al., 2025). Fourth, given that the clinical sample was composed mostly of chronic patients, it would be of interest to evaluate these findings in first-episode cohorts to assess their consistency in earlier phases of the disorder and reduce the possible influence of accumulated clinical factors. Finally, although the sample was restricted to individuals of European ancestry to ensure genomic homogeneity, previous evidence suggests that ubiquitination pathways are a cross-population risk factor (Liu et al., 2017). Therefore, our results should be confirmed in larger and more heterogeneous cohorts to evaluate the generalizability of these findings.

Taken together, our results reinforce the involvement of the ubiquitination system in the pathophysiology of schizophrenia, both in its etiological and cognitive dimensions. In particular, the greater relevance of USG subsets with prenatal and prefrontal expression suggests that this system’s contribution may be especially linked to early neurodevelopmental processes. Furthermore, the association between a higher polygenic load in USGs and poorer cognitive performance indicates that this pathway could contribute not only to disease risk but also to the cognitive heterogeneity observed among patients. Thus, the ubiquitination system emerges as a relevant component within the biological architecture of schizophrenia, and its integrated study at the genetic and functional levels may help to better understand the mechanisms linking genetic risk, neurodevelopment, and phenotypic variability in the disorder.

## Data Availability

The data supporting the findings of this study are available from the corresponding authors upon reasonable request.

## Author contributions

Conceptualization: G Riquelme-Alacid, B Ramos and M Fatjó-Vilas.

Data curation: G Riquelme-Alacid, M Guardiola-Ripoll, C Almodóvar-Payá.

Formal analysis: G Riquelme-Alacid.

Investigation: G Riquelme-Alacid, M Guardiola-Ripoll, D Herrera-Escartín, C Almodóvar-Payá,

N Hostalet, E Rodriguez-Cano, M Madre, E Pomarol-Clotet, B Ramos and M Fatjó-Vilas.

Methodology: G Riquelme-Alacid, M Guardiola-Ripoll, D Herrera-Escartín, B Ramos and M Fatjó-Vilas.

Visualization: G Riquelme-Alacid.

Resources: E Pomarol-Clotet, B Ramos and M Fatjó-Vilas.

Funding acquisition: M Madre, E Pomarol-Clotet, B Ramos and M Fatjó-Vilas.

Supervision: B Ramos and M Fatjó-Vilas.

Writing – original draft: G Riquelme-Alacid and M Fatjó-Vilas.

Writing – review & editing: All authors.

## Ethical Statement

Ethical approval was obtained from FIDMAG Hermanas Hospitalarias and Fundació Sant Joan de Déu Research Ethics Committees. All participants provided written informed consent about the study procedures and implications, which were carried out according to the Declaration of Helsinki and the ethical standards applicable to human research.

## Acknowledgements

The authors thank all the participants; without their generosity this study would not have been possible. This work has been carried out as part of the Bioquímica, Biología Molecular I Biomedicina doctoral programme at the Universitat Autònoma de Barcelona.

