## Supplementary methods, figures and tables for "Polygenic burden of ubiquitin system genes in schizophrenia: focus on prenatal neurodevelopment"

This document includes additional figures and tables supporting the results presented in the main manuscript. Supplementary figures include extended analyses of PGS models for ubiquitin system gene (USG) panels, as well as gene expression and enrichment profiles across tissues and developmental stages. Only significant results are included in the main figures; exploratory or complementary analyses are shown here. Figure and table numbering (e.g., Supplementary Figure 1, Supplementary Table 1, etc.) corresponds to citations in the main text and supplementary references.

**Table of Contents**


1. Supplementary Methods

1.1. Genomic DNA extraction, genotyping, quality control, and imputation.

1.2. Selection of ubiquitin system genes (USG).

1.3. Gene Set Analysis (GSA) of ubiquitin system genes using MAGMA.

1.4. Tissue and temporal expression profile of the gUSG panel.

1.5. Selection of USG panels based on spatiotemporal expression in brain development.

2. Supplementary Figures (included in this document)

2.1. Figure S2. Temporal expression and enrichment of gUSG panel.

2.2. Figure S3. Tissue specificity of SZ-associated in the gUSG.

2.3. Figure S4. Evaluation of polygenic score for USG panels using the clumping and thresholding method.

2.4. Figure S5. Competitive gene set analysis of ubiquitin system genes.

2.5. Figure S6. Discriminative performance of USG-based PGS models.

3. Supplementary Tables (included in this document)

3.1. Table S5. Full regression results for cognitive models (non-significant PGS associations).

3.2. Table S6. Regression models stratified by diagnostic group (SZ and HC): non-significant results.

4. Supplementary Reference

Provided in a separate Excel file (`Supplementary_Tables_file2.xlsx`)

Figure S1: Workflow for generating SNP panels for PGS analysis.

Table S1: GO terms filtered to define the ubiquitin system.

Table S2a: Global ubiquitin system genes (gUSG) panel.

Table S2b: SZ-associated SNPs mapped to gUSG.

Table S3a: Prenatal ubiquitin system genes (preUSG) panel.

Table S3b: Postnatal ubiquitin system genes (postUSG) panel.

Table S3c: Prenatal prefrontal cortex ubiquitin system genes (prePFC_USG) panel.

Table S3d: Postnatal prefrontal cortex ubiquitin system genes (postPFC-USG) panel.

Table S3e: Prenatal cerebellum ubiquitin system genes (preCB-USG) panel.

Table S3f: Postnatal cerebellum ubiquitin system genes (postCB-USG) panel.

Table S4a: LD-filtered SZ-associated SNPs in gUSG.

Table S4b: LD-filtered SZ-associated SNPs in preUSG.

Table S4c: LD-filtered SZ-associated SNPs in postUSG.

Table S4d: LD-filtered SZ-associated SNPs in prePFC_USG.

Table S4e: LD-filtered SZ-associated SNPs in postPFC_USG.

Table S4f: LD-filtered SZ-associated SNPs in preCB_USG.

Table S4g: LD-filtered SZ-associated SNPs in postCB_USG.

### 1. Supplementary Methods

*Supplementary Methods 1.1. Genomic DNA extraction, genotyping, quality control, and imputation*

Genomic DNA was obtained from all individuals using buccal mucosa swabs with the ATP Genomic DNA Mini Tissue Kit (Teknokroma Analítica, SA, Sant Cugat del Vallès, Barcelona, Spain) or from peripheral blood cells via venipuncture using the Realpure SSS Kit for DNA extraction (Durviz, SLU, Valencia, Spain).

Whole-genome genotyping was performed using the Infinium Global Screening Array-24 v1.0 (GSA) BeadChip (Illumina, Inc., San Diego, California, USA) at the Human Genotyping Laboratory of the Spanish National Cancer Research Centre (CeGen-ISCIII). A total of 730 059 SNPs were genotyped.

Quality control (QC) was performed using PLINK software versions 1.9 and 2.0 (Chang et al., 2015). SNPs were excluded if they deviated from Hardy-Weinberg equilibrium ($p < 1x10^{-6}$), had call rates < 98%, or minor allele frequency (MAF) < 0.005. At the sample level, individuals with missing data rates > 2% and those without European ancestry, as determined by principal component analysis, were excluded. After filtering, 447 035 SNPs and 315 individuals were retained.

Haplotype phasing was conducted using Eagle v2.4, and imputation was carried out with Minimac4, using the Haplotype Reference Consortium (HRC version r1.1) panel via the Michigan Imputation Server (https://imputationserver.sph.umich.edu/). Variants with MAF > 1% and imputation quality Rsq > 0.3 were retained. Genotypes were aligned to the GRCh37/hg19 human reference genome. The final post-imputation dataset included 7,606,397 genetic variants.

*Supplementary Methods 1.2. Selection of ubiquitin system genes (USG)*

Given the lack of a centralized reference for genes involved in the ubiquitin system, we applied a systematic approach based on Gene Ontology (GO) annotations (www.geneontology.org; (Carbon et al., 2019)). GO provides a structured and computationally accessible framework for annotating gene functions and biological processes.

The strategy was implemented in RStudio (R Studio Team, 2021; RStudio, 2011) and summarized in Supplementary Figure S1. Using the GO.db package (Carlson et al., 2019), we selected all GO terms whose name or definition contained the word "ubiquitin" (n=289). To refine specificity toward core ubiquitin-related processes, we excluded terms containing "ubiquitin-like", "sumo", "NEDD", "UFM", "no ubiquitin", and "ubiquitin independent", resulting in a curated set of 250 GO terms (see Supplementary Table S1).

To further characterize this set, the selected GO terms were manually categorized into four functional groups: ubiquitination (66.8%), deubiquitination (16.4%), substrate recognition (12%), and other associated processes (4.8%). Over 95% of the terms fell into the first three categories, indicating a high degree of specificity for core components of the ubiquitin system (see Supplementary Table S1).

Based on this curated and functionally categorized set of GO terms, we then extracted the corresponding human genes from the Ensembl database (GRCh37 assembly) using the biomaRt package (Durinck et al., 2009, 2005). Genes annotated with at least one of the selected GO terms yielded an initial list of 1,754 human genes. Genes located on sex chromosomes, alternative assemblies, or genomic patches, as well as genes lacking a valid match in GRCh37, were excluded. The final panel comprised 1,450 autosomal genes, which we defined as the global ubiquitin system genes (gUSG) (see Supplementary Table S2a).

*Supplementary Methods 1.3.* Gene Set Analysis (GSA) of ubiquitin system genes using MAGMA

Gene-set enrichment analysis was performed using MAGMA v1.10 (de Leeuw et al., 2015). Summary statistics from the schizophrenia GWAS (Trubetskoy et al., 2022) were harmonized with the 1000 Genomes Phase 3 European reference panel (GRCh37/hg19).

SNP-to-gene annotation was conducted using a window of 35 kb upstream and 10 kb downstream of gene boundaries. The extended major histocompatibility complex region (MHC; chromosome 6: 25–34 Mb) was excluded due to its complex linkage disequilibrium structure. Only autosomal genes included in the curated gUSG panel were retained.

Gene-level association statistics were computed by aggregating the signal from SNPs assigned to each gene while incorporating LD structure through the multiple regression framework implemented in MAGMA (de Leeuw et al., 2015), using the European 1000 Genomes panel as reference. Competitive gene-set analysis tested whether genes in the gUSG panel showed stronger associations with schizophrenia than other genes across the genome, adjusting for gene size, SNP density, minor allele count, and inter-gene LD, following the standard MAGMA implementation (de Leeuw et al., 2015).

To determine whether the observed enrichment exceeded chance expectations, 1,000 random gene sets were generated and matched to the gUSG panel based on (i) number of genes and (ii) SNP density distribution (Wang et al., 2011). This match preserved the genomic architecture of the panel and minimized potential inflation due to gene length or annotation biases. Each matched set was analyzed using the same annotation and gene-modelling pipeline applied to the gUSG panel.

*Supplementary Methods 1.4. Temporal and tissue expression profile of the gUSG panel*

To characterize the tissue and temporal expression profile of the gUSG panel, we conducted in silico analyses using the FUMA platform (http://fuma.ctglab.nl). We followed two complementary approaches: (1) temporal differential gene expression analysis, and (2) tissue enrichment analysis based on SZ-associated genetic variants located within the gUSG panel.

*1.4.1 Temporal expression of gUSG*

Temporal expression patterns of gUSG genes during human brain development were evaluated using the GENE2FUNC module of FUMA (Watanabe et al., 2017), which incorporates the BrainSpan dataset (Kang et al., 2011). This dataset includes both 11 major developmental periods and 29 more finely resolved developmental stages.

Genes were considered expressed when their RPKM (Reads Per Kilobase per Million mapped reads) was in at least 10% of the samples. Enrichment of differentially expressed genes at specific time points was assessed using a binomial test with Bonferroni correction ($p_{bon} < 0.05$), requiring a minimum of two overlapping genes per time point.

*1.4.2 Tissue enrichment of SZ-associated SNPs located in gUSG*

We further explored whether SZ-associated SNPs located within gUSG panel showed evidence of tissue-specific enrichment in brain regions. SNPs were extracted from the European subsample of the largest SZ GWAS to date (Trubetskoy et al., 2022). Using BEDTools v2.31.0 (Quinlan and Hall, 2010), we identified risk variants located within gUSG panel, without applying p-value filters to maximize the SNP set. We obtained a total of 220,128 SNPs (see Supplementary Table S2b).

The SNP set was analyzed using the SNP2GENE module of FUMA (Watanabe et al., 2017), with default parameters. Tissue enrichment analyses were conducted using MAGMA tool (de Leeuw et al., 2015), integrated into FUMA, using the GTEx v8 expression reference. Two GTEx panels were considered: a 30-tissue panel, which aggregates data from anatomically general tissue, and a 54-tissue panel, which includes more detailed information from specific anatomical regions (Aguet et al., 2019).

Genes were considered expressed in GTEx if their TPM (Transcripts Per Million) exceeded 0.1 in at least 10% of the sample. Associations were considered tissue-specific when $\beta > 0$ and $p < 0.001$ ($-\log(p) > 3$), following standard criteria.

In addition, we applied a more permissive exploratory threshold ($p < 0.00316$ ($-\log(p) > 2.5$) to capture moderate enrichment signals that, although not meeting strict significance threshold, may still reflect biologically relevant patterns. This approach was particularly relevant given the relatively restricted number of genes in the gUSG panel compared to genome-wide analyses, which may reduce statistical power. Applying a complementary threshold allowed us to identify consistent trends in tissues potentially relevant to SZ that might otherwise remain unperceived.

*Supplementary Method 1.5. Selection of USG panels based on spatiotemporal expression in brain development*

To explore whether the effects of USGs on SZ vulnerability might vary depending on their expression patterns during brain development, we generated six subsets from the gUSG panel using transcriptomic data from the Atlas of the Developing Human Brain (BrainSpan, www.brainspan.org) (see Supplementary Figure S1). This resource integrates RNA-seq and microarray data from postmortem human brain tissue of 57 donors spanning prenatal to adult stages (Kang et al., 2011; Sunkin et al., 2013).

*1.5.1 Selection of overexpressed USGs by developmental stage*

We used the differential expression tool provided by BrainSpan to compare gene expression between prenatal (8 – 38 post-conception weeks; pcw) and postnatal stages (birth – 40 years). Genes showing differential expression between these two developmental periods at $p < 5 \times 10^{-8}$ were retained, consistent with the genome-wide significance threshold commonly applied in large-scale genomic studies  (Pe'er et al., 2008). This resulted in 10,201 genes with higher prenatal expression and 9,826 with higher postnatal expression. These gene sets were intersected with the gUSG panel, generating two gene subsets: (1) preUSG panel: 654 USGs overexpressed in prenatal stages (see Supplementary Table 3a) and (2) postUSG panel: 357 USGs overexpressed in postnatal stages (see Supplementary Table 3b).

*1.5.2 Selection of USGs overexpressed in specific brain regions across developmental stages*

We applied the same comparison method to two brain regions identified as relevant in the tissue enrichment analysis: the prefrontal cortex (PFC) and the cerebellum (CB). For each structure, gene expression was compared between prenatal and postnatal stage using the same p-value threshold ($p < 5 \times 10^{-8}$). PFC included dorsolateral, ventrolateral, and anterior cingulate regions; CB included cerebellum and cerebellar cortex. Overlapping these region-specific produced four additional gUSG-derived regional subsets: (1) prePFC_USG: 477 USGs overexpressed in the PFC during prenatal stages (Supplementary Table S3c); (2) postPFC_USG: 208 USGs overexpressed in the PFC during postnatal stages (Supplementary Table S3d); (3) preCB_USG:

25 USGs overexpressed in the CB during prenatal stages (Supplementary Table S3e); and (4) postCB_USG: 130 USGs overexpressed in the CB during postnatal stages (see supplementary tables S3f).

### 2. Supplementary Figures

*2.1. Supplementary Figure S2*

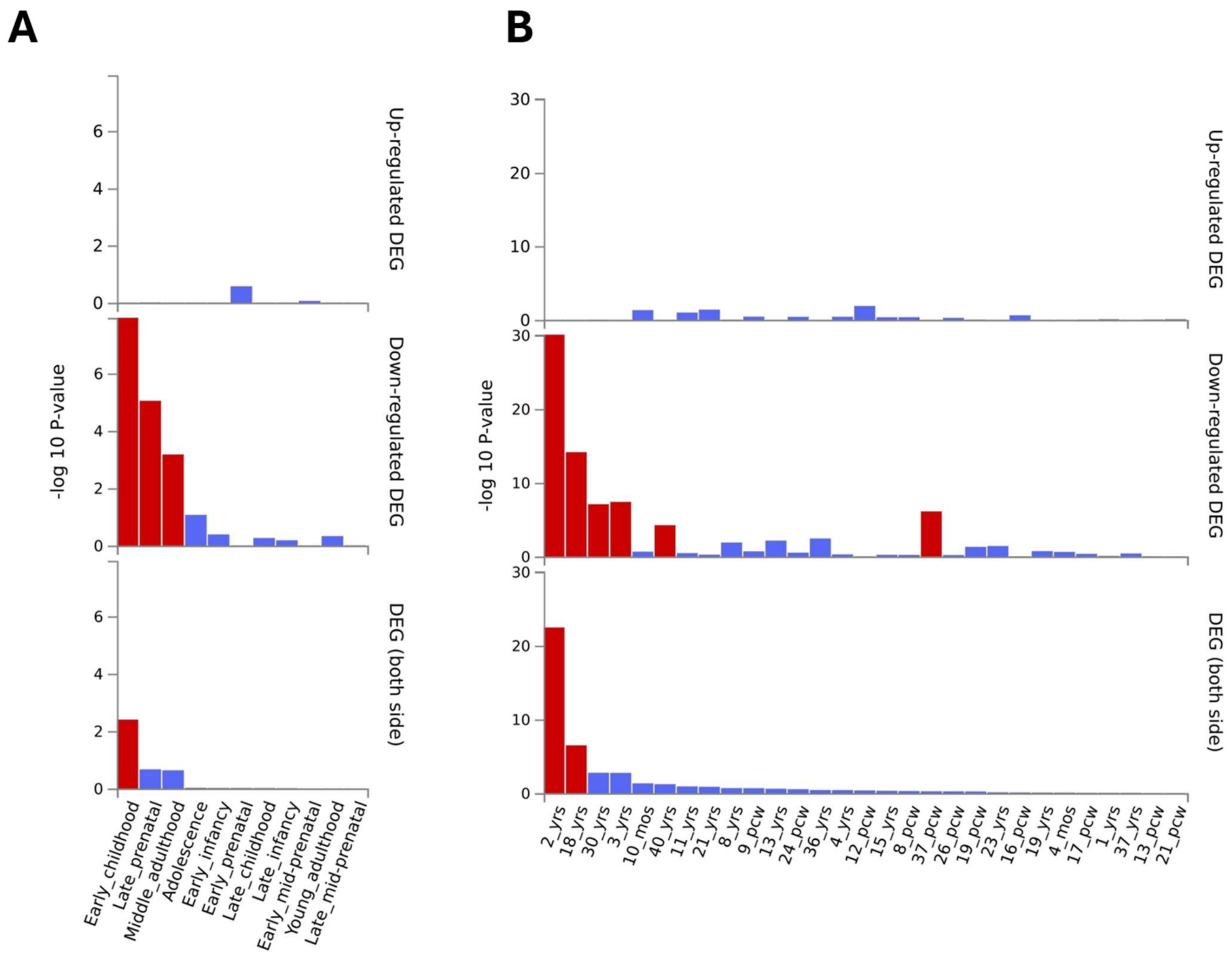


**Supplementary Figure S2. Temporal expression and enrichment of gUSG panel.** Spatiotemporal expression and enrichment profile of the gUSG panel in adult human tissue (GTEx v8) and across brain development (BrainSpan), obtained using the GENE2FUNC tool implemented in FUMA. **(A)**: Enrichment across 11 major developmental stages in the human brain (BrainSpan). **(B)**: Enrichment across 29 detailed chronological developmental stages (BrainSpan). Each panel includes three plots: Upper plot: Enrichment in genes with relatively high expression (up regulated) compared to the genomic background. Middle plot: Enrichment in genes with relatively low expression (down regulated). Lower plot: Overall enrichment overview sorted by statistical consistency across all stages/tissues. Highlights regions where gUSG overrepresentation is most robust (significant DEGs, Pbon < 0.05). Key findings: In BrainSpan (B), a sustained down-regulation pattern is observed during early childhood. In BrainSpan (B), this pattern persists throughout development, with significant enrichment particularly at 2 and 18 years of age. Notes: Red bars indicate significant enrichment (binomial test, Pbon < 0.05). Regulation direction reflects relative expression compared to the whole genome. Expression thresholds of TPM > 0.1 in GTEx and RPKM > 0.5 in BrainSpan were applied, required in at least 10% of samples.
*Abbreviations*: gUSG, global panel of ubiquitin system genes; DEG, differentially expressed gene; Pbon, Bonferroni-corrected p-value; TPM, transcripts per million; RPKM, reads per kilobase per million.

*2.2. Supplementary Figure S3*

**A**

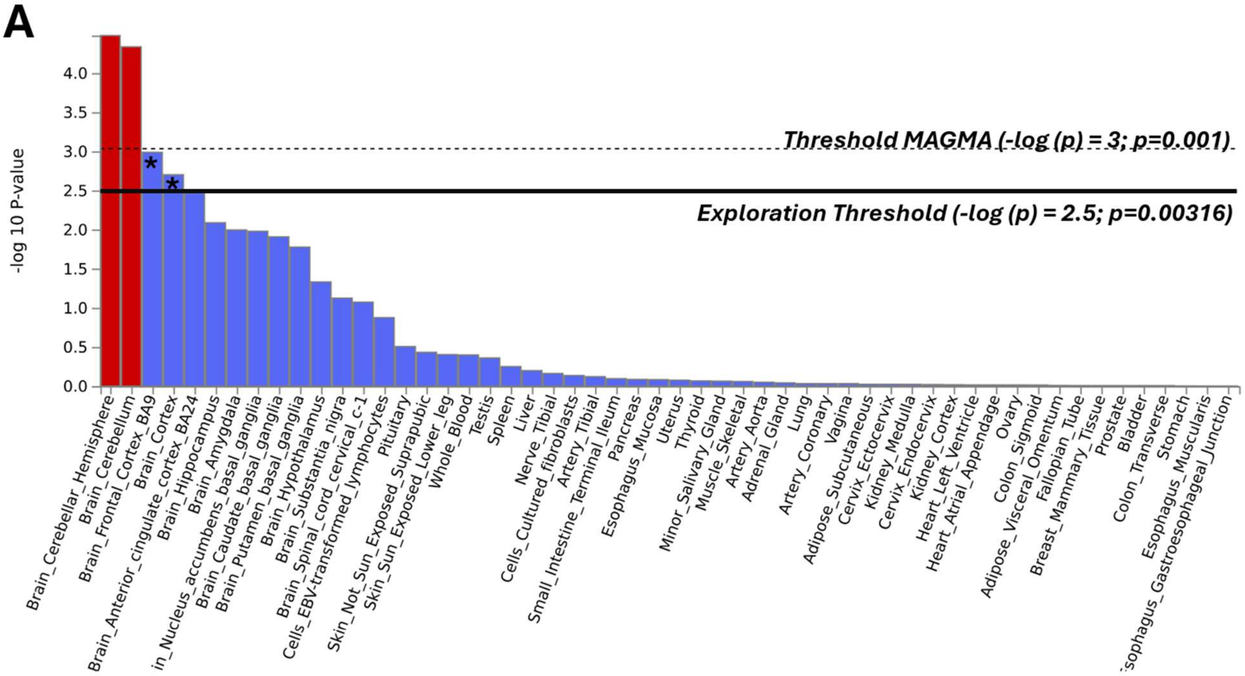


**B**

| FULL_NAME (tissue name) | BETA | P |
|---|---|---|
| Brain_Cerebellum | 0.13966 | **4.61E-05** |
| Brain_Cerebellar_Hemisphere | 0.13632 | **3.31E-05** |
| Brain_Frontal_Cortex_BA9 | 0.12843 | **0.0010305** |
| Brain_Cortex | 0.12336 | **0.0019892** |
| Brain_Anterior_cingulate_cortex_BA2 | 0.11516 | 0.0033892 |
| Brain_Hippocampus | 0.1116 | 0.0082004 |
| Brain_Caudate_basal_ganglia | 0.10555 | 0.012334 |
| Brain_Amygdala | 0.10478 | 0.010094 |
| Brain_Nucleus_accumbens_basal_ga | 0.10388 | 0.010498 |
| Brain_Putamen_basal_ganglia | 0.098724 | 0.016799 |
| Brain_Hypothalamus | 0.083387 | 0.046666 |
| Brain_Substantia_nigra | 0.07254 | 0.075305 |
| Brain_Spinal_cord_cervical_c-1 | 0.070951 | 0.084687 |
| Cells_EBV-transformed_lymphocytes | 0.030053 | 0.13352 |
| Pituitary | 0.02699 | 0.31444 |
| Skin_Not_Sun_Exposed_Suprapubic | 0.016484 | 0.37045 |
| Skin_Sun_Exposed_Lower_leg | 0.013403 | 0.3947 |
| Whole_Blood | 0.0078753 | 0.40078 |
| Testis | 0.0054849 | 0.43586 |

**Supplementary Figure S3. Tissue specificity of SZ-associated SNPs in the gUSG.** Tissue specificity analysis was performed using SNP2GENE tool implemented in FUMA, based on MAGMA, considering SNPs from the reference GWAS mapped to the gUSG. This analysis evaluates whether SZ-associated SNPs (according to summary statistics from the reference GWAS) are enriched in genes with high expression in specific tissues, adjusting for covariates such as gene size, SNP count and density, and mean gene expression (GTEx v8). **(A)**: Results of enrichment analysis for 54 specific tissues. Two reference thresholds are indicated: (i) Dashed line: default MAGMA significance threshold ($p < 0.001$; $-\log10(p) > 3$); and (ii) Solid black line: exploratory threshold adopted in this study ($p < 0.00316$; $-\log10(p) > 2.5$), used to prioritize tissues with potentially relevant signals, not meeting strict significance. Significant associations were found for the cerebellar hemisphere and cerebellum ($p < 0.001$; red bars); while brain FC BA9 and brain cortex were prioritized under the exploratory threshold ($p < 0.00316$; asterisks). **(B)**: Filtered view of panel B, displaying only tissues with a positive tissue specificity coefficient ($\beta > 0$) (those showing a positive association between gene expression and genetic signal). Green highlights indicate statistically significant tissues; yellow highlights indicate tissues prioritized under the exploration threshold; and gray highlights denotes tissues without relevant associations ($p > 0.00316$). β reflects the direction and strength of the association between gene expression in each tissue and genetic liability. The inclusion of an exploratory threshold allows for the consideration of moderate, potentially biologically relevant effects that may not survive multiple testing correction.

*Abbreviations*: gUSG, ubiquitin-proteasome system gene panel; MAGMA, Multi-marker Analysis of GenoMic Annotation; SZ, schizophrenia; SNP, single nucleotide polymorphism; p, probability value; β, tissue specificity coefficient; FC, frontal cortex.

#### *2.3. Supplementary Figure S4*

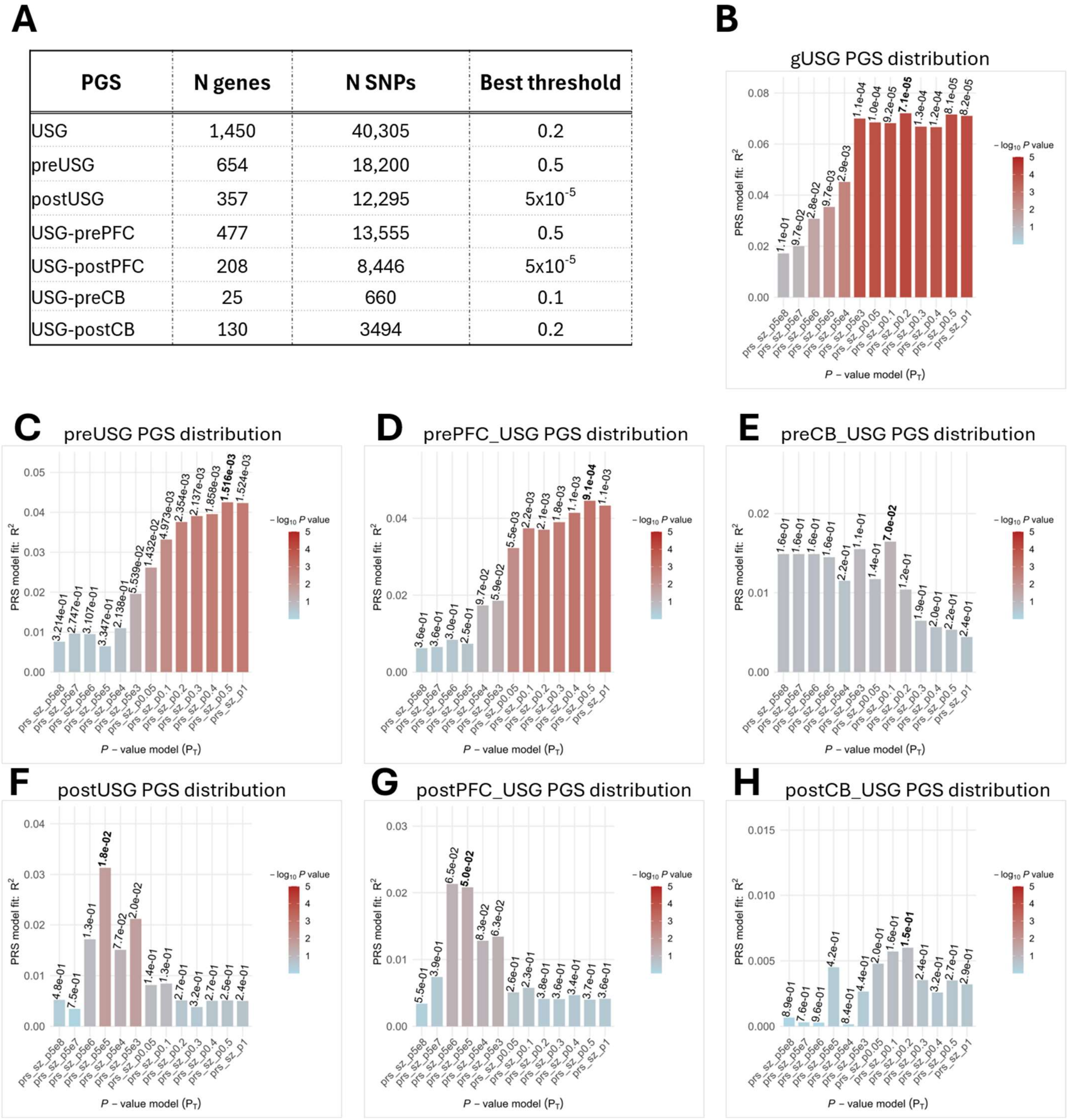

A

| PGS | N genes | N SNPs | Best threshold |
|---|---|---|---|
| USG | 1,450 | 40,305 | 0.2 |
| preUSG | 654 | 18,200 | 0.5 |
| postUSG | 357 | 12,295 | $5x10^{-5}$ |
| USG-prePFC | 477 | 13,555 | 0.5 |
| USG-postPFC | 208 | 8,446 | $5x10^{-5}$ |
| USG-preCB | 25 | 660 | 0.1 |
| USG-postCB | 130 | 3494 | 0.2 |



**Supplementary Figure S4. Evaluation of polygenic score for USG panels using the clumping and thresholding method.** **(A)** Summary table showing the numbers of genes and SNPs included in each PGS, the selected p-value threshold. **(B–H)** Bar plots showing Nagelkerke pseudo-$R^2$ across the evaluated p-value thresholds for each USG-derived PGS. Bar height represents model fit, and the corresponding model p-value is display above each bar. The selected threshold is identified by the bolded value. Panels correspond to **(B)** gUSG, **(C)** preUSG, **(D)** prePFC_USG, **(E)** preCB_USG, **(F)** postUSG, **(G)** postPFC_USG and **(H)** postCB_USG.

*Abbreviations*: PGS, polygenic score; USG, ubiquitination system genes; SNP, single-nucleotide polymorphism; C+T, clumping and thresholding; LD, linkage disequilibrium; gUSG, global USG; pre, prenatal; PFC, prefrontal cortex; CB, cerebellum; post, postnatal

*2.4. Supplementary Figure S5*

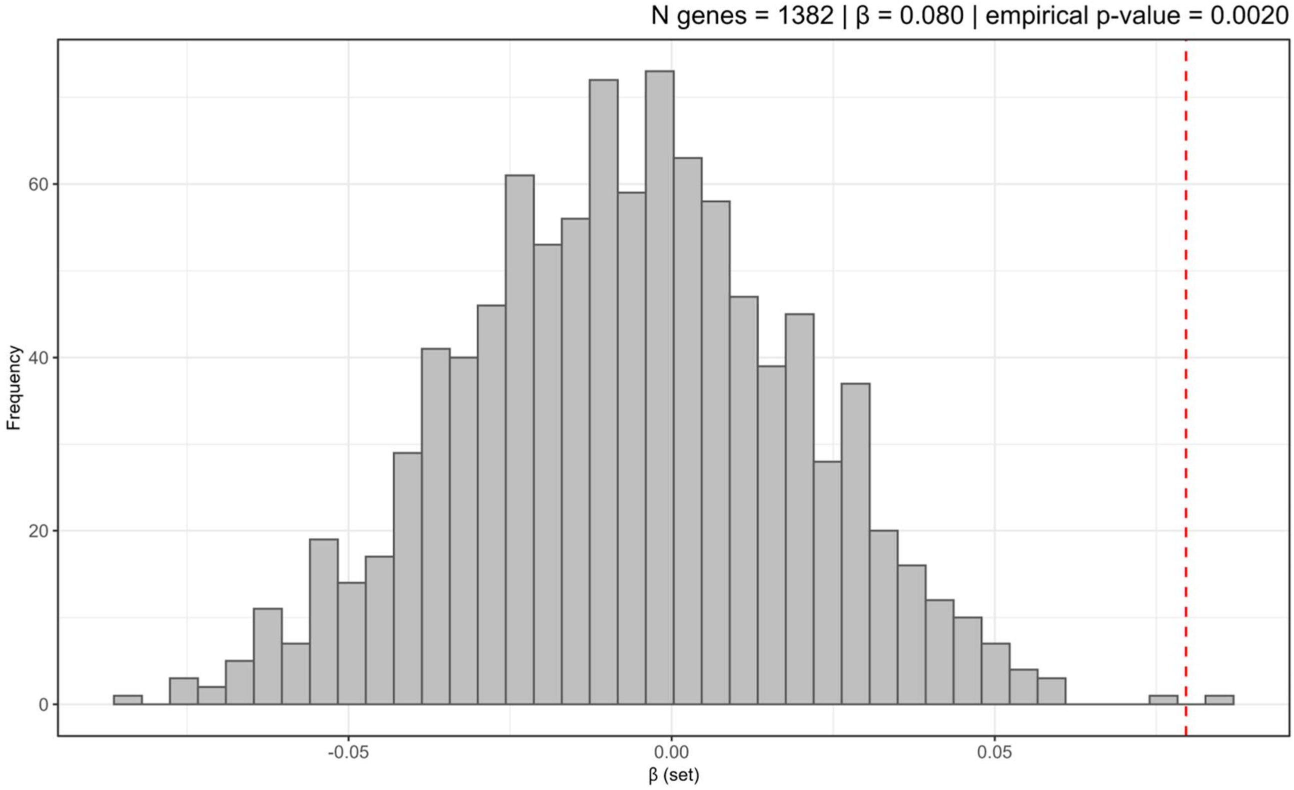


**Supplementary Figure S5. Competitive gene set analysis of ubiquitin system genes**. Distribution of effect sizes (β) obtained from 1,000 random gene sets matched to the global ubiquitin system gene panel (gUSG) in number of genes and SNP density. The dashed vertical line indicates the observed effect size of the gUSG panel (β = 0.080), which lies in the extreme right tail of the null distribution (empirical P = 0.002; 1 out of 1,000 null sets).

*2.5. Supplementary Figure S6*

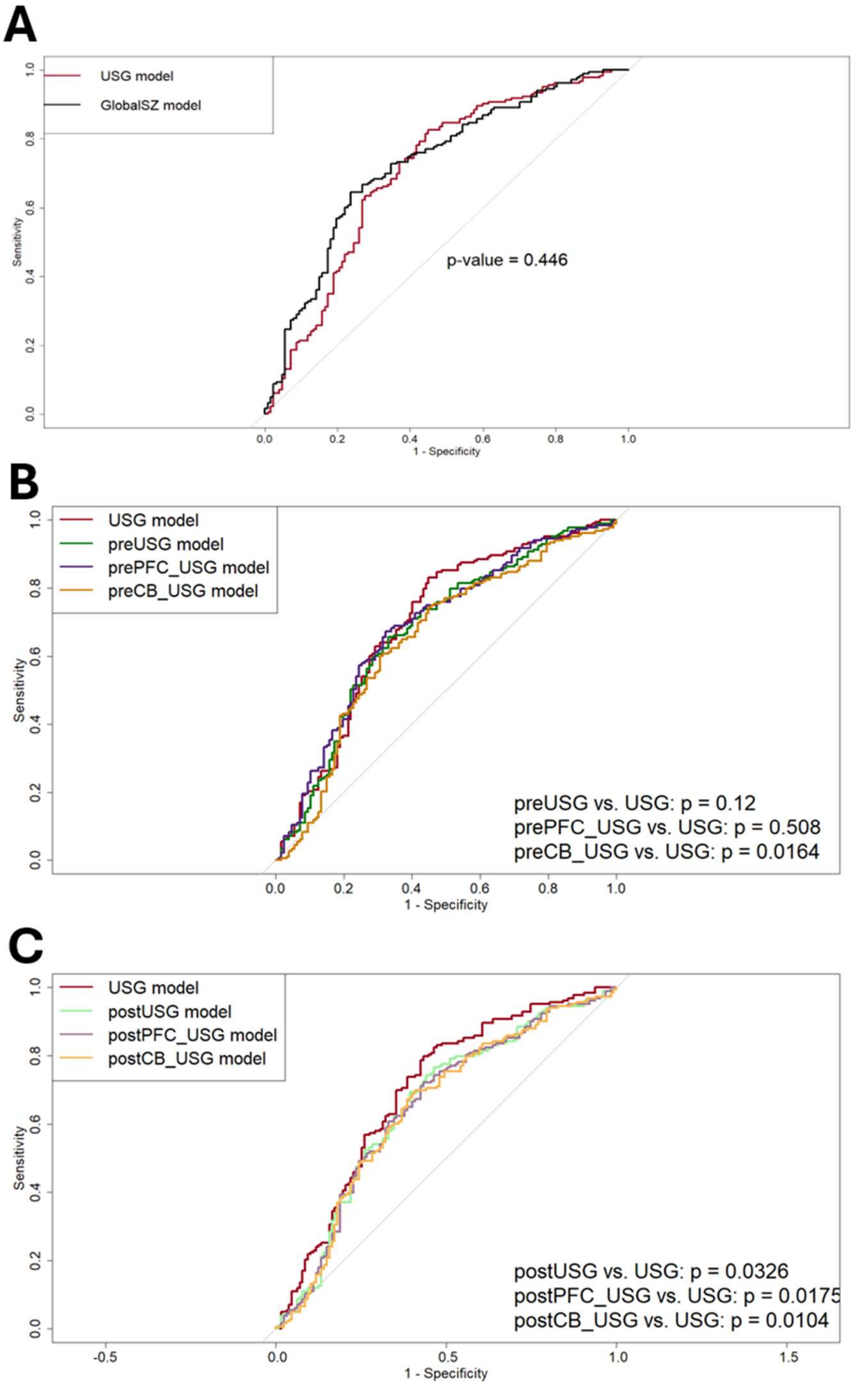


| PRS | AUC (95% C.I.) |
|---|---|
| USG | 0.71(0.65 - 0.77) |
| globalSZ | 0.73 (0.67 - 0.79) |

| PRS | AUC (95% C.I.) |
|---|---|
| preUSG | 0.68 (0.61 - 0.74) |
| prePFC_USG | 0.69(0.63 - 0.75) |
| preCB_USG | 0.65 (0.59 - 0.72) |

| PRS | AUC (95% C.I.) |
|---|---|
| postUSG | 0.66 (0.60 - 0.72) |
| postPFC_USG | 0.65 (0.59 - 0.71) |
| postCB_USG | 0.65 (0.58 - 0.71) |

**Supplementary Figure S6. Discriminative performance of USG-based PRS models.** (A) Comparison between the gUSG model and the globalSZ model using ROC curves.(B) Comparison between the gUSG model and USG panels defined by prenatal expression.(C) Comparison between the gUSG model and USG panels defined by postnatal expression. In each panel, ROC curves were generated using stratified repeated k-fold cross-validation (5 folds x 5 repetition), and p-values correspond to pairwise comparisons between the gUSG model and the respective models, calculated using DeLong's test for correlated ROC curves. The corresponding AUC values with 95% confidence intervals are presented in the adjacent tables.

Abbreviations: ROC, receiver operating characteristic; AUC, area under the curve; CI, confidence interval; gUSG, global USG panel; globalSZ, PRS derived from the GWAS reference for schizophrenia; pre, prenatal; PFC, prefrontal cortex; CB, cerebellum; post, postnatal.

### 3. Supplementary Tables

*3.1. Supplementary Table S5. Full regression results for cognitive models (non-significant PGS associations).*

| Outcome | PGS | Model summary | | Coefficients of the variable of interest (PGS) | | | | |
|---|---|---|---|---|---|---|---|---|
| | | adj. $R^2$ | $\Delta R^2$ | β (SE) | β est. | 95% I.C | p-value | $FDR_{pval}$ |
| TAP | USG | 0.12 | 0.006 | -0.59 (0.48) | -0.78 | [-1.54 – 0.36] | 0.224 | 0.299 |
| | preUSG | 0.12 | 0.007 | -0.98 (0.70) | -0.09 | [-2.36 – 040)] | 0.163 | 0.247 |
| | prePFC_USG | 0.11 | 0.008 | -1.16(0.83) | -0.09 | [-2.79 – 0.48] | 0.164 | 0.219 |
| WMS | USG | 0.43 | 0.001 | -0.23 (0.70) | -0.02 | [-1.61 – 1.15] | 0.574 | 0.574 |
| | preUSG | 0.43 | 0.001 | -0.45 (0.99) | -0.04 | [-2.41 – 1.52] | 0.651 | 0.651 |
| | prePFC_USG | 0.44 | 0.007 | -1.51 (1.28) | -0.09 | [-4.04 – 1.03] | 0.242 | 0.242 |

This table presents non-significant results from multiple linear regression models assessing the association between PGS and cognitive outcomes in intergroup analyses (including diagnosis as a covariate). All models were adjusted for age, sex, and the first four ancestry principal components. Results correspond to Block 2 (PGS effects). Nominal p-values < 0.05 are shown in bold for reference, although none survived false discovery rate (FDR) correction. Significant results are reported in Table 3 of the main manuscript.

*Abbreviations:* PGS, polygenic risk score; adj. $R^2$, adjusted R-squared; $\Delta R^2$, change in adjusted R-squared from Block 1; β (SE), unstandardized regression coefficient (standard error); β est., standardized regression coefficient; 95% CI, confidence interval of β; p-value and FDR p-value, nominal and false discovery rate–corrected p-values; USG, ubiquitin system genes; pre, prenatal; PFC, prefrontal cortex; TAP, Test de Acentuación de Palabras; BADS, Behavioural Assessment of the Dysexecutive Syndrome; WMS, Wechsler Memory Scale.

3.2. Table S6. Regression models stratified by diagnostic group (SZ and HC): non-significant results.

| SZ group | | | | | | | | |
|---|---|---|---|---|---|---|---|---|
| Outcome | PGS | Model summary | | Coefficients of the variable of interest (PGS) | | | | |
| | | adj. $R^2$ | $\Delta R^2$ | β (SE) | β est. | 95% I.C | p-value | $FDR_{pval}$ |
| TAP | preUSG | 0.04 | 0.025 | -1.90 (1.19) | -0.17 | [-4.26 – 0.46] | 0.113 | 0.145 |
| | prePFC_USG | 0.04 | 0.025 | -2.09 (1.32) | -0.16 | [-4.72 – 0.54] | 0.117 | 0.117 |
| WAIS-III | preUSG | 0.06 | 0.044 | -4.76 (2.26) | -0.22 | [-9.26 – (-0.257)] | **0.039** | 0.145 |
| | prePFC_USG | 0.05 | 0.036 | -5.09 (2.70) | -0.20 | [-10.47 – 0.28] | 0.063 | 0.084 |
| BADS | USG | 0.07 | 0.014 | -2.58 (2.91) | -0.13 | [-8.45 – 3.28] | 0.379 | 0.379 |
| | preUSG | 0.12 | 0.056 | -7.67 (4.27) | -0.26 | [-16.28 – 0.94] | 0.079 | 0.145 |
| | prePFC_USG | 0.15 | 0.076 | -10.8 (5.11) | -0.30 | [-21.17 – (-0.57)] | **0.039** | 0.084 |
| WMS | USG | 0.03 | 0.055 | -2.13 (1.28) | -0.26 | [-4.70 – 0.44] | 0.102 | 0.136 |
| | preUSG | 0.02 | 0.044 | -2.97 (1.99) | -0.25 | [-7.01 – 1.07] | 0.145 | 0.145 |
| HC group | | | | | | | | |
| Outcome | PGS | Model summary | | Coefficients of the variable of interest (PGS) | | | | |
| | | adj. $R^2$ | $\Delta R^2$ | β (SE) | β est. | 95% I.C | p-value | $FDR_{pval}$ |
| TAP | USG | 0.03 | 0.001 | 0.18 (0.60) | 0.03 | [-1.01 – 1.37] | 0.764 | ns |
| | preUSG | 0.03 | 0.000 | -0.04 (0.90) | -0.04 | [-1.83 –1.75] | 0.965 | ns |
| | prePFC_USG | 0.02 | 0.000 | 0.08 (1.12) | 0.01 | [-2.15 – 2.31] | 0.944 | ns |
| WAIS-III | USG | 0.07 | 0.022 | -1.73 (1.09) | -0.15 | [-3.90 – 0.433] | 0.116 | ns |
| | preUSG | 0.06 | 0.016 | -2.23 (1.67) | -0.13 | [-5.55 – 1.09] | 0.185 | ns |
| | prePFC_USG | 0.05 | 0.005 | -1.54 (2.09) | -0.07 | [-5.68 – 2.61] | 0.463 | ns |
| BADS | USG | 0.144 | 0.022 | -1.72 (1.37) | -0.15 | [-4.47 – 1.03] | 0.216 | ns |
| | preUSG | 0.155 | 0.032 | -2.91 (1.90) | -0.19 | [-6.72 – 0.90] | 0.132 | ns |
| | prePFC_USG | 0.155 | 0.032 | -3.77 (2.47) | -0.19 | [-8.71 – 1.18] | 0.133 | ns |
| WMS | USG | -0.04 | 0.021 | 1.00 (0.97) | 0.15 | [-0.95 – 2.96] | 0.307 | ns |
| | preUSG | -0.05 | 0.016 | 1.21 (1.36) | 0.13 | [-1.52 – 3.95] | 0.376 | ns |
| | prePFC_USG | -0.04 | 0.026 | 2.10 (1.81) | 0.17 | [-1.53 – 5.73] | 0.250 | ns |

This table presents non-significant results from multiple linear regression models assessing the association between polygenic risk scores (PGS) and cognitive outcomes in both the schizophrenia (SZ) and healthy control (HC) groups. All models were adjusted for age, sex, and the first four ancestry principal components. For the SZ group, models additionally included chlorpromazine (CPZ) equivalents and illness duration. Results correspond to Block 2 (PGS effects). Nominal p-values < 0.05 are shown in bold for reference, although none survived false discovery rate (FDR) correction. In the HC group, no p-values survived nominal significance; therefore, FDR correction was not applied. Significant associations are reported in Table 4 of the main manuscript.

*Abbreviations*: PGS, polygenic risk score; adj. $R^2$, adjusted R-squared; $\Delta R^2$, change in adjusted R-squared from Block 1; β (SE), unstandardized regression coefficient (standard error); β est., standardized regression coefficient; 95% CI, confidence interval of β; p-value and FDR p-value, nominal and false discovery rate–corrected p-values; USG, ubiquitin system genes; pre, prenatal; PFC, prefrontal cortex; TAP, Test de Acentuación de Palabras; WAIS-III, Wechsler Adult Intelligence Scale-Third Edition; BADS, Behavioural Assessment of the Dysexecutive Syndrome; WMS, Wechsler Memory Scale.

Carbon, S., Douglass, E., Dunn, N., Good, B., Harris, N.L., Lewis, S.E., Mungall, C.J., Basu, S., Chisholm, R.L., Dodson, R.J., Hartline, E., Fey, P., Thomas, P.D., Albou, L.P., Ebert, D., Kesling, M.J., Mi, H., Muruganujan, A., Huang, X., Poudel, S., Mushayahama, T., Hu, J.C., LaBonte, S.A., Siegele, D.A., Antonazzo, G., Attrill, H., Brown, N.H., Fexova, S., Garapati, P., Jones, T.E.M., Marygold, S.J., Millburn, G.H., Rey, A.J., Trovisco, V., Dos Santos, G., Emmert, D.B., Falls, K., Zhou, P., Goodman, J.L., Strelets, V.B., Thurmond, J., Courtot, M., Osumi, D.S., Parkinson, H., Roncaglia, P., Acencio, M.L., Kuiper, M., Lreid, A., Logie, C., Lovering, R.C., Huntley, R.P., Denny, P., Campbell, N.H., Kramarz, B., Acquaah, V., Ahmad, S.H., Chen, H., Rawson, J.H., Chibucos, M.C., Giglio, M., Nadendla, S., Tauber, R., Duesbury, M.J., Del, N.T., Meldal, B.H.M., Perfetto, L., Porras, P., Orchard, S., Shrivastava, A., Xie, Z., Chang, H.Y., Finn, R.D., Mitchell, A.L., Rawlings, N.D., Richardson, L., Sangrador-Vegas, A., Blake, J.A., Christie, K.R., Dolan, M.E., Drabkin, H.J., Hill, D.P., Ni, L., Sitnikov, D., Harris, M.A., Oliver, S.G., Rutherford, K., Wood, V., Hayles, J., Bahler, J., Lock, A., Bolton, E.R., De Pons, J., Dwinell, M., Hayman, G.T., Laulederkind, S.J.F., Shimoyama, M., Tutaj, M., Wang, S.J., D'Eustachio, P., Matthews, L., Balhoff, J.P., Aleksander, S.A., Binkley, G., Dunn, B.L., Cherry, J.M., Engel, S.R., Gondwe, F., Karra, K., MacPherson, K.A., Miyasato, S.R., Nash, R.S., Ng, P.C., Sheppard, T.K., Shrivatsav Vp, A., Simison, M., Skrzypek, M.S., Weng, S., Wong, E.D., Feuermann, M., Gaudet, P., Bakker, E., Berardini, T.Z., Reiser, L., Subramaniam, S., Huala, E., Arighi, C., Auchincloss, A., Axelsen, K., Argoud, G.P., Bateman, A., Bely, B., Blatter, M.C., Boutet, E., Breuza, L., Bridge, A., Britto, R., Bye-A-Jee, H., Casals-Casas, C., Coudert, E., Estreicher, A., Famiglietti, L., Garmiri, P., Georghiou, G., Gos, A., Gruaz-Gumowski, N., Hatton-Ellis, E., Hinz, U., Hulo, C., Ignatchenko, A., Jungo, F., Keller, G., Laiho, K., Lemercier, P., Lieberherr, D., Lussi, Y., Mac-Dougall, A., Magrane, M., Martin, M.J., Masson, P., Natale, D.A., Hyka, N.N., Pedruzzi, I., Pichler, K., Poux, S., Rivoire, C., Rodriguez-Lopez, M., Sawford, T., Speretta, E., Shypitsyna, A., Stutz, A., Sundaram, S., Tognolli, M., Tyagi, N., Warner, K., Zaru, R., Wu, C., Chan, J., Cho, J., Gao, S., Grove, C., Harrison, M.C., Howe, K., Lee, R., Mendel, J., Muller, H.M., Raciti, D., Van Auken, K., Berriman, M., Stein, L., Sternberg, P.W., Howe, D., Toro, S., Westerfield, M., 2019. The Gene Ontology Resource: 20 years and still GOing strong. Nucleic Acids Res. 47, D330–D338. https://doi.org/10.1093/nar/gky1055

Carlson, M., Falcon, S., Pages, H., Li, N., 2019. GO. db: A set of annotation maps describing the entire Gene Ontology. R Packag. version 3, 10–18129.

Chang, C.C., Chow, C.C., Tellier, L.C., Vattikuti, S., Purcell, S.M., Lee, J.J., 2015. Second-generation PLINK: rising to the challenge of larger and richer datasets. Gigascience 4, 7. https://doi.org/10.1186/s13742-015-0047-8

de Leeuw, C.A., Mooij, J.M., Heskes, T., Posthuma, D., 2015. MAGMA: generalized gene-set analysis of GWAS data. PLoS Comput. Biol. 11, e1004219. https://doi.org/10.1371/journal.pcbi.1004219

Durinck, S., Moreau, Y., Kasprzyk, A., Davis, S., De Moor, B., Brazma, A., Huber, W., 2005. BioMart and Bioconductor: a powerful link between biological databases and microarray data analysis. Bioinformatics 21, 3439–3440. https://doi.org/10.1093/bioinformatics/bti525

Durinck, S., Spellman, P.T., Birney, E., Huber, W., 2009. Mapping identifiers for the integration of genomic datasets with the R/Bioconductor package biomaRt. Nat. Protoc. 4, 1184–1191. https://doi.org/10.1038/nprot.2009.97

Kang, H.J., Kawasawa, Y.I., Cheng, F., Zhu, Y., Xu, X., Li, M., Sousa, A.M.M., Pletikos, M., Meyer, K.A., Sedmak, G., Guennel, T., Shin, Y., Johnson, M.B., Krsnik, Z., Mayer, S., Fertuzinhos, S., Umlauf, S., Lisgo, S.N., Vortmeyer, A., Weinberger, D.R., Mane, S., Hyde, T.M., Huttner, A., Reimers, M., Kleinman, J.E., Sestan, N., 2011. Spatio-temporal transcriptome of the human brain. Nature 478, 483–489. https://doi.org/10.1038/nature10523

Pe'er, I., Yelensky, R., Altshuler, D., Daly, M.J., 2008. Estimation of the multiple testing burden for genomewide association studies of nearly all common variants. Genet. Epidemiol. 32, 381–385. https://doi.org/10.1002/gepi.20303

Quinlan, A.R., Hall, I.M., 2010. BEDTools: a flexible suite of utilities for comparing genomic features. Bioinformatics 26, 841–842. https://doi.org/10.1093/bioinformatics/btq033

R Studio Team, 2021. A language and environment for statistical computing. R Found. Stat. Comput.

RStudio, 2011. RStudio: Integrated development environment for R (Version 0.97.311). J. Wildl. Manager.

Sunkin, S.M., Ng, L., Lau, C., Dolbeare, T., Gilbert, T.L., Thompson, C.L., Hawrylycz, M., Dang, C., 2013. Allen Brain Atlas: an integrated spatio-temporal portal for exploring the central nervous system. Nucleic Acids Res. 41, D996–D1008. https://doi.org/10.1093/nar/gks1042

Trubetskoy, V., Pardiñas, A.F., Qi, T., Panagiotaropoulou, G., Awasthi, S., Bigdeli, T.B., Bryois, J., Chen, C.-Y., Dennison, C.A., Hall, L.S., Lam, M., Watanabe, K., Frei, O., Ge, T., Harwood, J.C., Koopmans, F., Magnusson, S., Richards, A.L., Sidorenko, J., Wu, Y., Zeng, J., Grove, J., Kim, M., Li, Z., Voloudakis, G., Zhang, W., Adams, M., Agartz, I., Atkinson, E.G., Agerbo, E., Al Eissa, M., Albus, M., Alexander, M., Alizadeh, B.Z., Alptekin, K., Als, T.D., Amin, F., Arolt, V., Arrojo, M., Athanasiu, L., Azevedo, M.H., Bacanu, S.A., Bass, N.J., Begemann, M., Belliveau, R.A., Bene, J., Benyamin, B., Bergen, S.E., Blasi, G., Bobes, J., Bonassi, S., Braun, A., Bressan, R.A., Bromet, E.J., Bruggeman, R., Buckley, P.F., Buckner, R.L., Bybjerg-Grauholm, J., Cahn, W., Cairns, M.J., Calkins, M.E., Carr, V.J., Castle, D., Catts, S. V, Chambert, K.D., Chan, R.C.K., Chaumette, B., Cheng, W., Cheung, E.F.C., Chong, S.A., Cohen, D., Consoli, A., Cordeiro, Q., Costas, J., Curtis, C., Davidson, M., Davis, K.L., de Haan, L., Degenhardt, F., DeLisi, L.E., Demontis, D., Dickerson, F., Dikeos, D., Dinan, T., Djurovic, S., Duan, J., Ducci, G., Dudbridge, F., Eriksson, J.G., Fañanás, L., Faraone, S. V, Fiorentino, A., Forstner, A., Frank, J., Freimer, N.B., Fromer, M., Frustaci, A., Gadelha, A., Genovese, G., Gershon, E.S., Giannitelli, M., Giegling, I., Giusti-Rodríguez, P., Godard, S., Goldstein, J.I., González Peñas, J., González-Pinto, A., Gopal, S., Gratten, J., Green, M.F., Greenwood, T.A., Guillin, O., Gülöksüz, S., Gur, R.E., Gur, R.C., Gutiérrez, B., Hahn, E., Hakonarson, H., Haroutunian, V., Hartmann, A.M., Harvey, C., Hayward, C., Henskens, F.A., Herms, S., Hoffmann, P., Howrigan, D.P., Ikeda, M., Iyegbe, C., Joa, I., Julià, A., Kähler, A.K., Kam-Thong, T., Kamatani, Y., Karachanak-Yankova, S., Kebir, O., Keller, M.C., Kelly, B.J., Khrunin, A., Kim, S.-W., Klovins, J., Kondratiev, N., Konte, B., Kraft, J., Kubo, M., Kučinskas, V., Kučinskiene, Z.A., Kusumawardhani, A., Kuzelova-Ptackova, H., Landi, S., Lazzeroni, L.C., Lee, P.H., Legge, S.E., Lehrer, D.S., Lencer, R., Lerer, B., Li, M., Lieberman, J., Light, G.A., Limborska, S., Liu, C.-M., Lönnqvist, J., Loughland, C.M., Lubinski, J., Luykx, J.J., Lynham, A., Macek, M.J., Mackinnon, A., Magnusson, P.K.E., Maher, B.S., Maier, W., Malaspina, D., Mallet, J., Marder, S.R., Marsal, S., Martin, A.R., Martorell, L., Mattheisen, M., McCarley, R.W., McDonald, C., McGrath, J.J., Medeiros, H., Meier, S., Melegh, B., Melle, I., Mesholam-Gately, R.I., Metspalu, A., Michie, P.T., Milani, L., Milanova, V., Mitjans, M., Molden, E., Molina, E., Molto, M.D., Mondelli, V., Moreno, C., Morley, C.P., Muntané, G., Murphy, K.C., Myin-Germeys, I., Nenadić, I., Nestadt, G., Nikitina-Zake, L., Noto, C., Nuechterlein, K.H., O'Brien, N.L., O'Neill, F.A., Oh, S.-Y., Olincy, A., Ota, V.K., Pantelis, C., Papadimitriou, G.N., Parellada, M., Paunio, T., Pellegrino, R., Periyasamy, S., Perkins, D.O., Pfuhlmann, B., Pietiläinen, O., Pimm, J., Porteous, D., Powell, J., Quattrone, D., Quested, D., Radant, A.D., Rampino, A., Rapaport, M.H., Rautanen, A., Reichenberg, A., Roe, C., Roffman, J.L., Roth, J., Rothermundt, M., Rutten, B.P.F., Saker-Delye, S., Salomaa, V., Sanjuan, J., Santoro, M.L., Savitz, A., Schall, U., Scott, R.J., Seidman, L.J., Sharp, S.I., Shi, J., Siever, L.J., Sigurdsson, E., Sim, K., Skarabis, N., Slominsky, P., So, H.-C., Sobell, J.L., Söderman, E., Stain, H.J., Steen, N.E., Steixner-Kumar, A.A., Stögmann, E., Stone, W.S., Straub, R.E., Streit, F., Strengman, E., Stroup, T.S., Subramaniam, M., Sugar, C.A., Suvisaari, J., Svrakic, D.M., Swerdlow, N.R., Szatkiewicz, J.P., Ta, T.M.T., Takahashi, A., Terao, C., Thibaut, F., Toncheva, D., Tooney, P.A., Torretta, S., Tosato, S., Tura, G.B., Turetsky, B.I., Üçok, A., Vaaler, A., van Amelsvoort, T., van Winkel, R., Veijola, J., Waddington, J., Walter, H., Waterreus, A., Webb, B.T., Weiser, M., Williams, N.M., Witt, S.H., Wormley, B.K., Wu, J.Q., Xu, Z., Yolken, R., Zai, C.C., Zhou, W., Zhu, F., Zimprich, F., Atbaşoğlu, E.C., Ayub, M., Benner, C., Bertolino, A., Black, D.W., Bray, N.J., Breen, G., Buccola, N.G., Byerley, W.F., Chen, W.J., Cloninger, C.R., Crespo-Facorro, B., Donohoe, G., Freedman, R., Galletly, C., Gandal, M.J., Gennarelli, M., Hougaard, D.M., Hwu, H.-G., Jablensky, A. V, McCarroll, S.A., Moran, J.L., Mors, O., Mortensen, P.B., Müller-Myhsok, B., Neil, A.L., Nordentoft, M., Pato, M.T., Petryshen, T.L., Pirinen, M., Pulver, A.E., Schulze, T.G., Silverman, J.M., Smoller, J.W., Stahl, E.A., Tsuang, D.W., Vilella, E., Wang, S.-H., Xu, S., Adolfsson, R., Arango, C., Baune, B.T., Belangero, S.I., Børglum, A.D., Braff, D., Bramon, E., Buxbaum, J.D., Campion, D., Cervilla, J.A., Cichon, S., Collier, D.A., Corvin, A., Curtis, D., Forti, M. Di, Domenici, E., Ehrenreich, H., Escott-Price, V., Esko, T., Fanous, A.H., Gareeva, A., Gawlik, M., Gejman, P. V, Gill, M., Glatt, S.J., Golimbet, V., Hong, K.S., Hultman, C.M., Hyman, S.E., Iwata, N., Jönsson, E.G., Kahn, R.S., Kennedy, J.L., Khusnutdinova, E., Kirov, G., Knowles, J.A., Krebs, M.-O., Laurent-Levinson, C., Lee, J., Lencz, T., Levinson, D.F., Li, Q.S., Liu, J., Malhotra, A.K., Malhotra, D., McIntosh, A., McQuillin, A., Menezes, P.R., Morgan, V.A., Morris, D.W., Mowry, B.J., Murray, R.M., Nimgaonkar, V., Nöthen, M.M., Ophoff, R.A., Paciga, S.A., Palotie, A., Pato, C.N., Qin, S., Rietschel,

M., Riley, B.P., Rivera, M., Rujescu, D., Saka, M.C., Sanders, A.R., Schwab, S.G., Serretti, A., Sham, P.C., Shi, Y., St Clair, D., Stefánsson, H., Stefansson, K., Tsuang, M.T., van Os, J., Vawter, M.P., Weinberger, D.R., Werge, T., Wildenauer, D.B., Yu, X., Yue, W., Holmans, P.A., Pocklington, A.J., Roussos, P., Vassos, E., Verhage, M., Visscher, P.M., Yang, J., Posthuma, D., Andreassen, O.A., Kendler, K.S., Owen, M.J., Wray, N.R., Daly, M.J., Huang, H., Neale, B.M., Sullivan, P.F., Ripke, S., Walters, J.T.R., O'Donovan, M.C., 2022. Mapping genomic loci implicates genes and synaptic biology in schizophrenia. Nature 604, 502–508. https://doi.org/10.1038/s41586-022-04434-5

Wang, L., Jia, P., Wolfinger, R.D., Chen, X., Zhao, Z., 2011. Gene set analysis of genome-wide association studies: methodological issues and perspectives. Genomics 98, 1–8. https://doi.org/10.1016/j.ygeno.2011.04.006

Watanabe, K., Taskesen, E., van Bochoven, A., Posthuma, D., 2017. Functional mapping and annotation of genetic associations with FUMA. Nat. Commun. 8, 1826. https://doi.org/10.1038/s41467-017-01261-5
